# Elevated levels of environmental enteric dysfunction biomarkers among rural Indonesian infants: associations with water, sanitation, hygiene and linear growth

**DOI:** 10.64898/2026.02.19.26346361

**Authors:** Callum Lowe, Tony Arjuna, Mubasyir Hasanbasri, Haribondhu Sarma, I Nyoman Sutarsa, Severine Navarro, Darren Gray, Matthew Kelly

## Abstract

**Objective:** To investigate the burden of environmental enteric dysfunction (EED) and its association with water, sanitation, and hygiene (WASH) and linear growth amongst infants in rural Central Java, Indonesia.

**Study design:** A longitudinal study of 119 infants aged between 5-19 months was conducted in five villages of Wonosobo District, Central Java, Indonesia. Anthropometric measurements of infants and their mothers were performed at baseline and 5-month follow-up alongside a quantitative questionnaire on household, socio-economic, WASH and caregiving variables and stool sample collection for the investigation of alpha-1-antitrypsin (AAT), neopterin (NEO), and myeloperoxidase (MPO) levels. Linear mixed-effects regression models estimated the associations between WASH and height-for-age z-score (HAZ) on log-transformed EED biomarkers.

**Results:** Biomarkers increased from baseline to follow-up despite a declining trend with age and 68.7%, 79.0%, and 71.4% of infants experienced elevated AAT, NEO, and MPO respectively follow-up. Infants had higher AAT if they averaged > 30 minutes playing on soiled surfaces per day (β = 0.11, p<0.05). NEO was elevated in infants with diarrhoea (β = 1.04, p<0.05), municipal water source ( = β 0.71, p<0.05), and in infants who mouthed soiled fomites weekly (β = 0.55, p<0.05). Infants in houses with municipal water source had higher MPO (β = 0.56, p<0.05) and higher MPO if mouthing soil weekly (β = 0.41, p<0.05). Compared to infants at risk of stunting, stunted infants at baseline had lower AAT at follow-up (β = −0.39, p<0.05) while infants with HAZ > −1 had lower AAT at baseline (β = −0.43, p<0.05). HAZ at baseline was positively associated with NEO at follow-up (β = 0.36, p<0.05). MPO was higher in infants with HAZ > −1 at follow-up (β = 0.59, p<0.05) and stunted infants (β = −0.54, p<0.05) compared to infants at risk of stunting.

**Conclusion:** Elevated EED biomarker levels were frequent and associated weakly with WASH and HAZ with bi-directionality, highlighting the need for quality birth cohort studies to improve understanding of EED and develop interventions.

## Introduction

Poor water, sanitation, and hygiene (WASH) conditions are prevalent in low– and middle-income countries (LMICs) and remain a pervasive global issue (1). Nearly one billion people practice open defecation and do not have access to improved water resulting in a chronic exposure to gastrointestinal pathogens with significant health and economic impacts in affected communities (2). These conditions can lead to vicious cycle of acute malnutrition and diarrhoea, the latter of which claims the lives of one million people, mainly children, annually (3). Poor WASH conditions, in addition to non-infectious causes such as diets low in dietary diversity and animal-source foods, are also a major driver of chronic undernutrition, or stunting, which affects at least 149 million children under the age of five years globally (4, 5). Stunted children on average have reduced earnings as an adult (6), impaired cognitive development (7) and for girls increased risk of birth complications and the risk of giving birth to stunted children (8).

WASH interventions in LMICs have failed to demonstrate noticeable impacts on linear growth (9, 10). While many plausible reasons for this exist including methodological issues such as poor intervention implementation, adherence and compliance, a major point of concern is the probability that household WASH interventions fail to break known and underrecognized exposure and transmission pathways that are unique to babies (11). The degree of exposure is profound, with infants in low-resource settings demonstrated to have higher levels of faecal bacteria on their hands compared to their high-income counterparts (12). High exposure to faecal bacteria has been noted also in infants complementary foods, caregivers hands, play objects and in soil ingested (“geophagia”) as part of exploratory play behaviour (13). This chronic, high exposure to faecal pathogens has been shown to lead to structural and physiological changes to the small intestine (14). These changes, including but not limited to villous blunting, crypt hyperplasia, intestinal inflammation and translocation of bacterial products across a damaged intestinal barrier into systemic inflammation has been referred to as environmental enteric dysfunction (EED) (15). EED has been proposed as a missing link in understanding the causal chain between poor WASH conditions and linear growth faltering amongst infants in LMICs (16). The lack of impact of household WASH interventions on EED further supports the hypothesis that reducing chronic exposure to faecal pathogens that drive EED is critical to achieve reductions in stunting (17, 18).

EED is increasingly viewed as a chronic mucosal adaptation to repeated early-life enteric pathogen exposure in settings of poverty, food insecurity and suboptimal WASH, rather than a single-pathogen syndrome (19). Recurrent microbial and dietary insults drive epithelial stress and impaired barrier function (tight-junction disruption, altered production of mucus and anti-microbial peptides), with increased permeability and luminal antigen (pathogen-associated molecular patterns) access to the lamina propria (20). This promotes sustained innate immune activation, such as neutrophils and activation of macrophages and monocytes, and downstream inflammatory remodelling of the small intestine, including villus blunting, crypt hyperplasia and altered absorption and metabolic activity (21). Whilst histology of small intestine biopsy samples remains the gold standard to demonstrate epithelial injury with heightened mucosal activation to formally diagnose EED; serum, urine and stool biomarkers of one or more domains of EED has become the main approach to investigating EED (22). Major strides have been made in identifying biomarkers of EED such as stool alpha-1-antitrypsin to assess protein loss/permeability, myeloperoxidase representative of neutrophil activation, neopterin for IFN-γ associated macrophage activation and serum biomarkers such as lipopolysaccharide and endotoxin core antibody (23), and some studies have found associations between these biomarkers and associated low-grade enteropathy can reduce nutrient absorption and increase systemic inflammation likely contributing to poor linear growth and immune development amongst infants in LMICs (24, 25). There are some limited findings related to associations between EED biomarkers and infant-play behaviours such as geophagia amongst infants in Bangladesh (26, 27) as well as close living proximity to domesticated livestock (28). Overall, however, the body of evidence linking infant-specific risk factors to EED is limited and there is a need for research to identify risk factors as they may be potential targets of ‘baby’ WASH interventions (16).

In addition to this, the association between biomarkers of EED and linear growth is likely complex and non-linear (29). Biomarkers may exhibit seasonality (30) and levels of biomarkers may be reduced due to downregulation of the immune system amongst infants who have experienced an exhaustive load of pathogen exposure (31, 32). This could lead to bidirectionality where EED initially drives poor linear growth that then shapes subsequent immune responses towards recurrent faecal pathogens. The interpretation of high and low may be blurred or ambiguous if this were the case.

Indonesia is a middle-income archipelago nation of 283 million residents, of which 100 million do not have access to improved sanitation and 117 million do not have access to improved water (33). Regions in Indonesia with worse WASH conditions have higher rates of child stunting (34). Whilst improvements have been made since 2018 in reducing child stunting, prevalence remains high in rural areas (35) and nationally is at 19.8% in 2024 (36). Indonesia’s climate exhibits relatively definitive wet and dry seasons which may add an additional layer of complexity to understanding EED (30). Stunting interventions in Indonesia are often limited to nutritional supplementation (37) which has marginal impact on linear growth (38). There is little research on EED in south-east Asia, particularly Indonesia (31). However, existing research has investigated EED in poor areas where basic sanitation infrastructure and piped water are missing (39) as well as densely populated urban slums (40, 41). It remains relatively unexplored however as to whether EED is prevalent and/or a driver of stunting in communities that have basic WASH infrastructure but persistently high stunting rates. Considering these knowledge gaps and areas for further investigation, we conducted a longitudinal study of infants in rural Indonesia with the aim of measuring the association between WASH risk factors with EED biomarkers and linear growth faltering. We also explored interactions with age and season to improve the understanding of nuances in the measurement of EED biomarkers and their trends over time.

## Methods

This study was reported in line with the Strengthening the Reporting of Observational Studies in Epidemiology (STROBE) statement (Supplementary Table 8) (42).

### Study design, sample and population

This was a longitudinal observational study of rural infants from five villages in Wonosobo, Central Java, Indonesia. This is a mountainous area, and study villages were located between 5-20 kilometres from the Wonosobo town centre with household income predominantly arising from farming and livestock, small/micro businesses and manual labour jobs. Wonosobo is the district with the highest stunting prevalence in Central Java province (43).

Data were collected at two points – baseline data was collected during the dry season between August – September 2024, follow-up data was collected five months later during the wet season between January – February 2025. The wet and dry seasons in this mountainous area are not as contrasted as in other areas of Indonesia but are noticeable as fewer days of rain and lighter rain. The target age group for infants was six to eighteen months at baseline as this is the period of most significant growth faltering (44).

Infants were selected by multi-stage random sampling. Originally, two districts were selected in Wonosobo and a list of villages (“*Desa*”) across the two districts was obtained. Two villages in the smaller district and three villages in the larger district were randomly selected, and a list of all infants in each village combined with their sex, mothers name, and household address was requested from district public health centres (“*Puskesmas*”). This list was narrowed down to infants who would be aged between 6– and 18-months at the time of planned baseline data collection. The number of infants recruited from each village was proportional to the total number of infants in each village.

Due to limited information on the central tendency and dispersion of EED biomarkers particularly amongst Indonesian infants, the sample size was determined with the Cohen’s d calculation as one sufficient to detect a priori 50% effect size in EED biomarker concentrations between infants with HAZ < −1 and HAZ ≥ −1 with 5% probability of a type I error and 80% power which requires 128 infants. Due to restrictions in the size of ELISA test kits sold, the sample size had to be reduced to 120 which equates to a reduction in power from 80% to 77%.

Criteria for infants inclusion were that they were not suffering from a known chronic disease or severe infection requiring treatment (e.g. malaria, tuberculosis, dengue) and whose mother was willing to provide informed consent. Infants were excluded if their family had not been living in the current place of residence for the past 3-months or planned to move in the upcoming 6-months.

### Data collection

Prior to data collection, a training event was held to train data collectors on administration of the questionnaire, stool sample collection procedures, and anthropometric measurements of infants and mothers. Data collectors were local research assistants with prior experience in similar fieldwork. Data was regularly cross-checked between data collectors. Re-training of data collectors was performed prior to follow-up.

Data was collected via two household visits at baseline and at follow-up. Data collection at baseline involved a quantitative questionnaire with infants’ mothers using REDCap electronic data capture on tablets, anthropometric measurements of infants and their mother, and stool sample collection. Follow-up data collection was similar except for mothers’ anthropometric measurements omitted and utilised a shortened questionnaire only consisting of time-varying exposures and covariates.

The quantitative questionnaire pertained to household demographic and socio-economic factors, key maternal and paternal covariates, ownership of livestock, proximity of livestock to the household, infants’ health and diet, infants play activities, exposure to livestock, and key behaviours that may increase the risk of EED (e.g. frequency of geophagia, playing on a soiled surface). Further detail is provided in the exposures section. Consumption of animal-source foods (ASF) was measured through a simple food frequency questionnaire as it is a key predictor of linear growth (45) and we theorise low intake may increase susceptibility to EED. Mothers were requested to respond to how frequently their infants had been consuming ASF as multiple times per day, daily, weekly, monthly, or never. These were added up across different categories to obtain the overall estimated daily serves of ASF.

Infant length was recorded using an infant length board (InnoQ Infantometer 100, PT Astra Components Indonesia, 1mm graduation), and weight using a calibrated baby weight scale (InnoQ Digital Baby Scale 100, PT Astra Components Indonesia, 5g graduation). For one participant, weight was measured as the average of two deducted weights (mother’s/local health cadre’s weight while cradling infant to chest minus own weight). Infants wore minimal clothing during weight measurement. Maternal height was measured using a portable stadiometer (InnoQ Stadiometer 100, 1mm graduation) and weight with a standing weight scale (InnoQ Digital Flat Scale 100, 50g graduation). No infants required their length (height) measured standing.

### Stool sample collection and analysis

At baseline and follow-up, mothers of infants were requested to assist in providing a non-diarrhoeal stool sample for the analysis of EED biomarkers. For infants who were experiencing diarrhoea at the time of data collection, mothers were requested to not collect any stool until infants’ faeces returned to normal texture and consistency. Mothers were provided with a stool sample collection container with an internally attached spatula, gloves, and zip-lock plastic and an additional plastic bag. Mothers were requested to collect a grape-sized amount of stool from infants’ diapers and to aim to collect stool from multiple areas in the diaper if possible. Stool samples were kept at 2-8°C for <24 hours before being transported to a local laboratory for storage at –20°C for up to a month before being transported to a central laboratory in Jakarta and stored at –70°C prior to biomarker analysis.

Stool samples were analysed to determine the concentration of faecal myeloperoxidase (MPO), neopterin (NEO), and alpha-1-antitrypsin (AAT). Analysis was performed using commercial ELISA kits according to manufacturer instructions (AAT & MPO: Immundiagnostik AG, Bensheim, Germany, NEO: IBL Immuno-biological laboratories, Minnesota USA). Samples were diluted 1:25,000 for AAT, 1:8 for NEO, and 1:500 for MPO. Produced in the liver, AAT is an enzyme inhibitor that protects the lungs from enzymes, however its presence in the stool is an indicator of protein leakage in the intestinal tract (46), a hallmark of EED due to damage of the tight junctions between epithelial cells (47). NEO is produced by macrophages after interferon-gamma stimulation indicating intestinal immune activity and inflammation (48). MPO is primarily produced by neutrophils and is involved in the production of antimicrobial reactive oxygen species, therefore, it is an indicator of intestinal immune activity and inflammation. High MPO production is also damaging to normal intestinal tissue (49).

### WASH variables

A broad set of WASH exposures were considered in the quantitative questionnaire so to capture multiple potential pathways of faecal pathogen exposure and important covariates. Four major categories were developed: 1) household WASH conditions (water source for cooking and drinking, improved sanitation, unsafe disposal of infant faeces, and presence of dirt floors in the household), 2) exposure to livestock faeces (poultry, cat, and bird faeces), 3) caregiver WASH (storage of infants food at room temperature between feedings, mother washes hands with soap before preparing food, father handles animals and/or animal faeces), and 4) play behaviour risks (infant regularly plays in gardens, infant regularly plays on roads, average daily time infant spends playing directly on soil/dirt/sand surfaces, frequency infant mouths soil/sand/dirt – “geophagia”, frequency infant mouths soiled or dirty fomites (e.g. outdoor play items, hand tools), frequency infant mouths animal faeces).

### Statistical analysis

#### Data preparation

Data cleaning was performed in IBM SPSS Version 26 (50) and all analysis performed in RStudio (51). Anthropometry z-scores were computed in WHO AnthroPlus software (52) which has the 2006 WHO Child Growth Charts built-in (53). No outliers were detected using the cutoffs of +/− 6 standard deviations. Significant associations were p < 0.05 and borderline associations were reported if p < 0.10.

#### Descriptive and bivariate analysis

Descriptive analysis was performed to tabulate counts of infants across demographic, health, and WASH variables. The mean and standard deviation of anthropometric z-scores was calculated for baseline and follow-up. Median and interquartile range of EED biomarkers was calculated for baseline and follow-up ant the proportion of observations at each time point that were elevated was calculated using the following cut-offs commonly used in EED literature: AAT [<27 mg/dL], NEO [<70 nmol/L], MPO [<2000 ng/mL] (54). Pearson correlations were computed for biomarkers within and between time points. Student’s t-tests were computed to compare the mean of log-transformed biomarkers across four key health variables: high ASF consumption (≥1.5 serves per day vs < 1.5 as a general proxy of higher consumption without detailed food volume data), recent 7-day diarrhoea, 3-month fever, and current breastfeeding status. Scatter plots of the age-biomarker relationship at each time point were overlayed with a simple linear model to visualise the impact of seasonality considering a hypothesized declining trend of the biomarkers with age. Kernel density plots were produced to visualize the raw distribution of biomarkers across boys and girls.

#### Association between WASH and EED biomarkers

To account for repeated biomarker measurements within children and for clustering of children within villages, we fitted a linear mixed effects model with random intercepts for infant and village. This approach allowed pooling of baseline and follow-up data and estimating interactions with time. Although some predictors were time-invariant (e.g., SES characteristics), their effects on biomarker concentrations could vary over time. This was important to investigate because biomarkers showed low correlation within children across time points, indicating that both time-varying exposures and time-varying effects of stable predictors contribute to the observed variation. The models were developed in an iterative process:

Base mode: A null model with only random intercepts for infant and village was fitted to quantify the proportion of total variability attributed to clustering.

Step 1: A priori infant-level covariates were added: age, sex, time, current breastfeeding (yes/no), mother’s age, animal-source food intake, recent (7-day) diarrhoea, 3-month fever, and time of day of bowel motion of the stool sample provided. Each variable was tested for an interaction with time and significant interactions (p<0.05) were retained. This set reflects key demographic and health determinants expected to influence biomarker concentrations.

Step 2 (Socioeconomic model): Core SES confounders (mothers’ highest education (senior secondary or higher vs. lower), household expenses (> Rp. 2.000.000 per month vs below), and fathers’ occupation (employee/government vs. manual labour/farmer/small business owner) were included a priori regardless of statistical significance. Additional SES indicators (household bedroom count, ownership of livestock, wall material, and ownership of household assets including fridge, Wi-Fi, water heater) were included only if their main effect or time interaction reached p<0.05. Average monthly household expenses were selected over household income as expenses are more reflective of long-term average income as month-to-month income can vary greatly in these rural areas.

Step 3 (WASH models): WASH variables were added into the socioeconomic model with three separate models. The first model was a theory-based model only including WASH variables (water source, household sanitation, presence of dirt floors in the household, and unsafe disposal of infant faeces). An interaction time was included if statistically significant. The second (exploratory) model consisted of all the WASH and infant play variables captured in the quantitative questionnaire including their interaction with time. To mitigate multicollinearity, WASH variables with VIF > 5 in a full model were excluded before selection. Remaining variables and their time interactions were selected using hybrid stepwise selection (entry/removal cut-off p<0.01). This model was used to identify under-recognised WASH-EED pathways relevant to the local setting. For the third model, a WASH index was created where variables within each conceptual domain were summed to create four binary high vs. low-risk indices (cut-offs chosen to ensure > 25% of infants were in the high-risk group). The four indices were modelled together without individual WASH variables. Where interactions with time were significant at p<0.10, we use the emmeans package in R to extract the baseline– and follow-up specific effects. Anthropometry was not included in any of these models to prevent conditioning on a collider which could lead to distorted associations between WASH and EED biomarkers.

#### Association between attained HAZ and EED biomarkers

Although EED is commonly conceptualised as a cause of impaired growth, the association is likely bidirectional; impaired linear growth may reflect chronic immune activation and influence subsequent intestinal immune activation and permeability. To model how infants in different linear growth states differ in their biomarker concentrations, we modelled HAZ as an exposure in the same socioeconomic-adjusted linear mixed effects framework used for the WASH-EED analysis including significant WASH predictors of HAZ.

HAZ was examined both as a continuous predictor and as a categorical variable to compare biomarker levels at both time points between infants with higher attained linear growth (HAZ > −1), infants at risk of stunting (−2 < HAZ ≤ −1) and infants who were stunted (HAZ ≤ −2). WHZ was included in all models to account for potential effects of acute malnutrition on biomarker concentrations. Anthropometry was modelled in two ways. In the first, HAZ and WHZ were treated as time-varying predictors to model baseline HAZ/WHZ with baseline biomarkers and follow-up HAZ/WHZ with follow-up biomarkers. This represents the cross-sectional instantaneous association at each time point; that is *“What is the instantaneous association between an infants attained height-for-age or weight-for-height on their current state of intestinal inflammation and intestinal permeability?*”. Time interactions were used to test whether the association differed between baseline and follow-up. The second way anthropometry was modelled was the temporal effect by explicitly modelling baseline HAZ/WHZ together with a time interaction, allowing estimation of whether attained growth at baseline predicted biomarker concentrations at follow-up. This can be thought of as “*What is the effect of an infant’s height-for-age or height-for-weight on their intestinal inflammation and permeability in the future?*”. We did not model the relationship between follow-up HAZ explicitly with baseline biomarkers because there was no rationale, but the relationship between baseline biomarkers and subsequent growth faltering (dHAZ) was modelled and described in the next section.

#### Association between EED and growth faltering

The effect of EED on linear growth faltering was measured using ordinary least squares linear regression models with ΔHAZ as the dependent variable adjusting for baseline HAZ to account for regression to the mean. The models were adjusted for an a priori list of key covariates and confounders for linear growth including age, sex, breastfeeding status, birthweight, mother’s height, mothers age, mother’s education, household expenses, fathers’ occupation, animal-source food intake, diarrhoea, fever, and significant WASH predictors of ΔHAZ. Models were fitted for baseline and follow-up biomarkers separately to avoid overfitting, and within each time point all biomarkers were modelled together to estimate their isolated effects on ΔHAZ. Baseline biomarkers regressed onto ΔHAZ reflect a temporal relationship, reflecting whether early EED predicted subsequent growth faltering. Associations between follow-up biomarkers and ΔHAZ may reflect whether ongoing or recent EED occurring during the follow-up period in which ΔHAZ was measured. For comparability with other methods in the literature, the biomarkers were modelled as tertile comparisons (above 75^th^ percentile and below 25^th^ percentile vs. 25^th^—75^th^ percentile). We explored whether there the role of EED in driving growth faltering differs depending on baseline attained height through an interaction term between biomarkers and the HAZ_BL_ in the linear model of ΔHAZ.

### Ethics

This project received ethical approval from the Indonesian National Research and Innovation Agency (BRIN) Health Research Ethics Committee (Protocol #26032024004917) and the Australian National University Research Ethics Committee (Protocol H/2023/1123). Mothers provided informed consent for their participation in the research project after reading an information sheet outlining the project goals, involvement, risks and benefits. Mothers could withdraw from the project at any time with no implications.

## Results

### Descriptive statistics of the study population

One hundred and twenty (120) participants were recruited at baseline with one lost to follow-up, leading to a total sample of 119 infants with non-diarrhoeal stool samples provided at baseline and follow-up. All proceeding analysis is presented for these 119 infants. Participants were evenly split by sex (47.1% female) and across age groups (Table 1). Due to birthdate reporting errors in local data systems, infants age at baseline was between five and nineteen months instead of the intended six to eighteen months. Most infants were from households with average monthly expenses between 1 – 2 million rupiah (approx. $60 – $120 USD) and approximately a third (32.8%) of households had average monthly expenses above 2 million rupiah. Just over a half of households (52.9%) owned livestock. Just above half of households owned a fridge (55.8%) and fewer owned Wi-Fi (38.3%) and fewer owned a water heater (21.7%). Most households (81.5%) had painted walls.

**Table 1.**
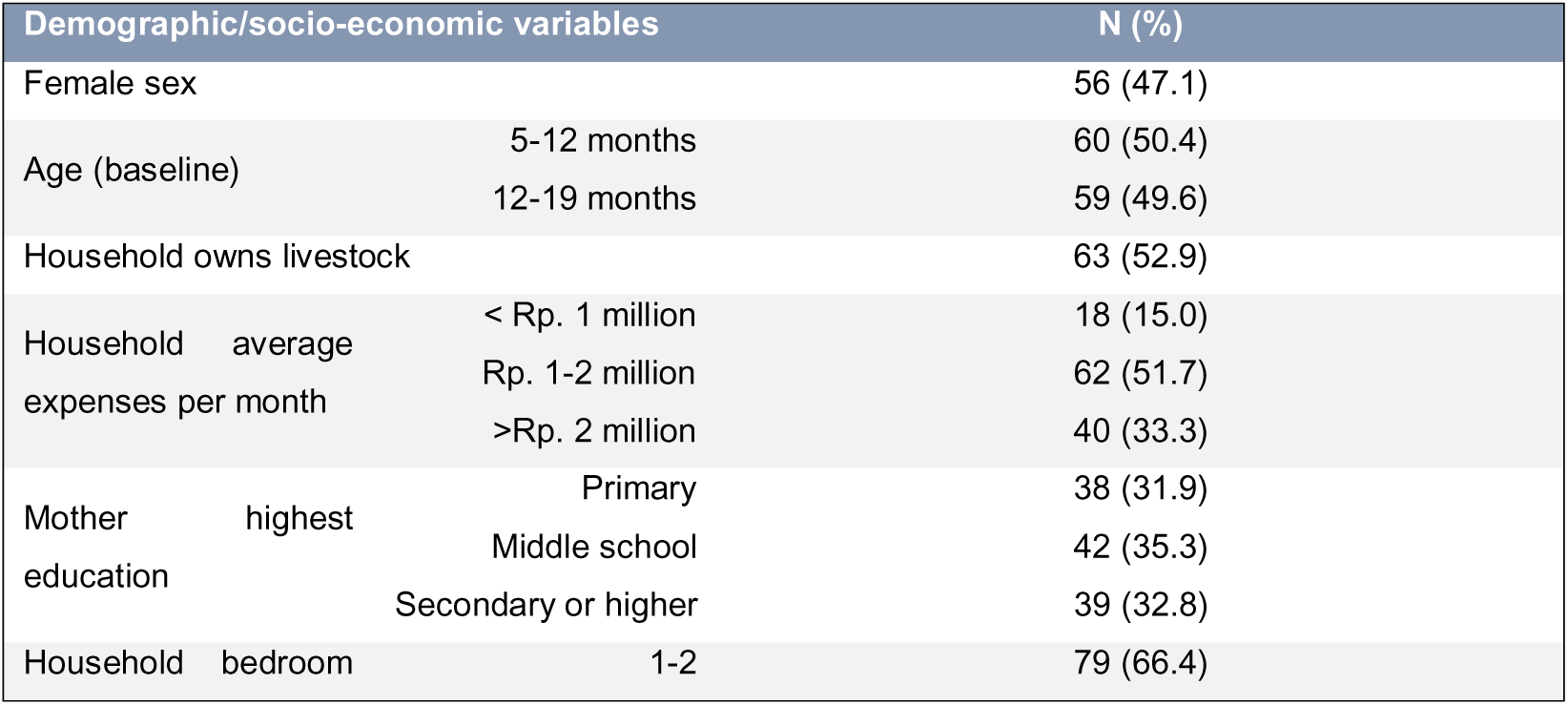
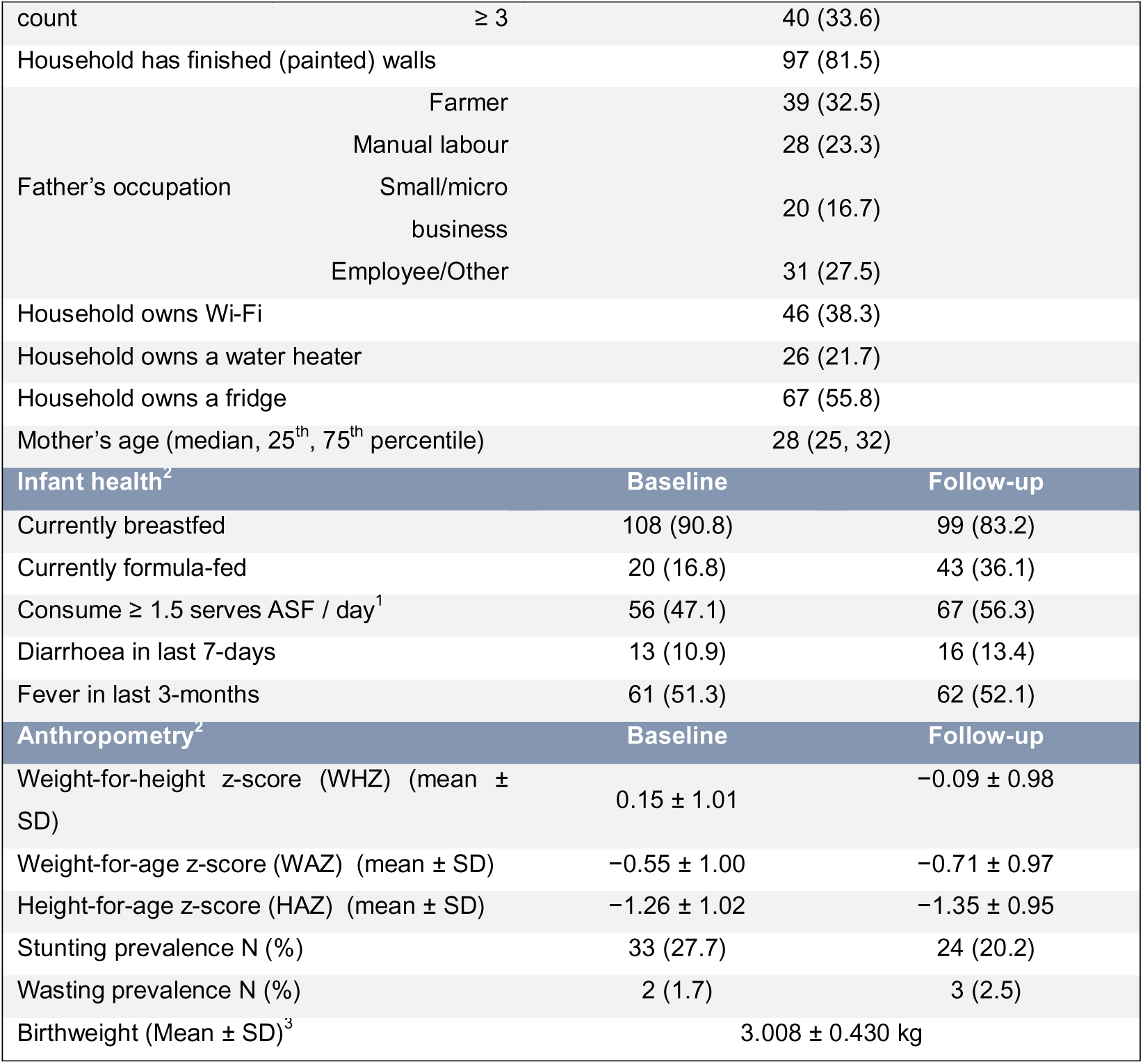
Demographic, socio-economic, health and anthropometry of the study sample. Diarrhoea and fever were self-reported by infants’ mothers. Stunting defined as HAZ ≤−2, wasting as WHZ ≤−2. ^1^Consumption of ASF (animal source foods) obtained from a food frequency questionnaire. **^2^**Infant health and anthropometry were measured at baseline and at follow-up while demographic and socioeconomic variables were recorded at baseline only. **^3^**Birthweights were recorded from local vital health data books.

| Demographic/socio-economic variables |  |  | N (%) |
| --- | --- | --- | --- |
| Female sex |  |  | 56 (47.1) |
| Age (baseline) | 5-12 months |  | 60 (50.4) |
|  | 12-19 months |  | 59 (49.6) |
| Household owns livestock |  |  | 63 (52.9) |
| Household average expenses per month | < Rp. 1 million |  | 18 (15.0) |
|  | Rp. 1-2 million |  | 62 (51.7) |
|  | >Rp. 2 million |  | 40 (33.3) |
| Mother highest education | Primary |  | 38 (31.9) |
|  | Middle school |  | 42 (35.3) |
|  | Secondary or higher |  | 39 (32.8) |
| Household bedroom | 1-2 |  | 79 (66.4) |

**Table 1.** Demographic, socio-economic, health and anthropometry of the study sample.
|  |  |  |
| --- | --- | --- |
| count | ≥ 3 | 40 (33.6) |
| Household has finished (painted) walls |  | 97 (81.5) |
| Father's occupation | Farmer | 39 (32.5) |
|  | Manual labour | 28 (23.3) |
|  | Small/micro business | 20 (16.7) |
|  | Employee/Other | 31 (27.5) |
| Household owns Wi-Fi |  | 46 (38.3) |
| Household owns a water heater |  | 26 (21.7) |
| Household owns a fridge |  | 67 (55.8) |
| Mother's age (median, 25<sup>th</sup>, 75<sup>th</sup> percentile) |  | 28 (25, 32) |
| **Infant health<sup>2</sup>** |  |  |
|  | **Baseline** | **Follow-up** |
| Currently breastfed | 108 (90.8) | 99 (83.2) |
| Currently formula-fed | 20 (16.8) | 43 (36.1) |
| Consume ≥ 1.5 serves ASF / day<sup>1</sup> | 56 (47.1) | 67 (56.3) |
| Diarrhoea in last 7-days | 13 (10.9) | 16 (13.4) |
| Fever in last 3-months | 61 (51.3) | 62 (52.1) |
| **Anthropometry<sup>2</sup>** |  |  |
|  | **Baseline** | **Follow-up** |
| Weight-for-height z-score (WHZ) (mean ± SD) | 0.15 ± 1.01 | -0.09 ± 0.98 |
| Weight-for-age z-score (WAZ) (mean ± SD) | -0.55 ± 1.00 | -0.71 ± 0.97 |
| Height-for-age z-score (HAZ) (mean ± SD) | -1.26 ± 1.02 | -1.35 ± 0.95 |
| Stunting prevalence N (%) | 33 (27.7) | 24 (20.2) |
| Wasting prevalence N (%) | 2 (1.7) | 3 (2.5) |
| Birthweight (Mean ± SD)<sup>3</sup> | 3.008 ± 0.430 kg |  |

Common occupations of infants’ fathers were farmer (32.5%), manual labour jobs (23.3%) and owner of a small or micro business (16.7%). Mothers’ education was low with 32.8% of mothers reaching secondary high school or higher education. Most infants were currently being breastfed at baseline (90.8%) which only dropped slightly to 83.2% at follow-up. Approximately one third of infants (36.1%) were currently receiving formula milk at follow-up. About half of the infants consumed ≥ 1.5 serves of animal-sourced food per day at both baseline and follow-up. Seven-day diarrhoea prevalence increased from 10.9% at baseline to 13.4% at follow-up. Approximately half of mothers at baseline (51.3%) and follow-up (52.1%) reported their infant had a fever in the last three months.

Infants were born with birth weight on average 3,008g and only three infants were wasted at follow-up. Stunting prevalence was slightly above a quarter (27.7%) at baseline but mean HAZ was low at −1.26 at baseline which decreased to −1.35 by follow-up, corresponding to an average ΔHAZ of −0.09. The loss of WAZ over the follow-up period was larger dropping from −0.55 to −0.71 at follow-up which corresponded with a drop in WHZ from 0.15 at baseline to −0.09 at follow-up.

### WASH conditions and infant play behaviour

Overall, WASH conditions were mixed between participants households (Table 2). At the household level, WASH risk factors were prevalent with partial dirt floors (29.4% of households), unimproved sanitation (37.0% of households), unsafe disposal of infant faeces (30.2% of households). Unimproved sanitation among sample households was exclusively toilets piped to a fishpond or river.

**Table 2.** WASH conditions in the study sample. Data expressed as N (%). ^1^Merged cells are for time-independent variables captured in the baseline questionnaire only. ^2^Data expressed as median (25^th^, 75^th^ percentile)

| **Household WASH conditions<sup>1</sup>** |  |
| --- | --- |
| Spring water source (vs. municipal) | 84 (70.6) |
| Unimproved sanitation | 44 (37.0) |
| Infant faeces disposed unsafely | 36 (30.2) |
| Partial dirt/earth floors (vs. fully cemented/tiled) | 35 (29.4) |
| Father handles livestock or livestock faeces | 44 (36.7) |
| **Mother's WASH<sup>1</sup>** |  |
| Mother washes hands with soap before preparing meals | 78 (65.5) |
| Infants' foods are stored at room temperature for prolonged periods | 59 (49.6) |

**Table 2.** WASH conditions in the study sample.
| Exposure to livestock | Baseline | Follow-up |
| --- | --- | --- |
| Exposed to poultry faeces | 10 (8.4) | 16 (13.4) |
| Exposed to bird faeces | 16 (13.4) | 25 (21.0) |
| Exposed to cat faeces | 35 (29.4) | 48 (40.3) |
| Play behaviour |  |  |
| Mouth soil/sand/dirt at least weekly | 12 (10.1) | 39 (32.8) |
| Mouth soiled fomites at least weekly | 28 (23.5) | 67 (56.3) |
| Ever mouth animal faeces (vs. never) | 5 (4.2) | 5 (4.2) |
| Average daily play time on soiled surfaces<sup>2</sup> | 10 (0, 35) | 30 (30, 60) |
| Infant plays on nearby roads | 27 (22.6) | 29 (24.4) |
| Infant plays in nearby gardens | 22 (18.5) | 24 (20.2) |

Food and water risks were present with municipal piped tap water provided to only 29.4% of infants’ households while 70.6% relied on piped water from nearby springs, and infant foods were routinely stored at room temperature for prolonged periods between feedings by 49.6% of mothers. Livestock faeces exposure was not commonly reported with only 13.4% of infants exposed to poultry faeces and 21.0% exposed to bird faeces, although 40.3% were exposed to cat faeces and 36.7% of infants had a father who regularly handled livestock or livestock faeces. 65.5% of mothers reported washing their hands with soap before preparing meals.

Mothers reported their infants to generally play and engage in exploratory behaviour in their environments which was more prevalent at follow-up. Mouthing of soil/sand/dirt, or geophagia, was practiced at least weekly by 32.7% of infants at follow-up which was an increase from 10.1% at baseline. More frequently however was the mouthing of soiled fomites which increased from 23.6% at least weekly at baseline to 56.3% at least weekly at follow-up. Mouthing of animal faeces was rare and only reported by the mothers of five infants at baseline. A smaller proportion of mothers reported their infants regularly played on nearby roads (24.4%) or gardens (20.2%) at follow-up. The median of the average daily time infants spent playing on soiled surfaces as reported by mothers increased from 10 minutes at baseline to 30 minutes at follow-up.

### Crude biomarker levels

EED biomarkers were elevated at baseline and follow-up in most infants (Supplementary Table 6). The median (interquartile range) of AAT at baseline was 37.2 (21.4 – 70.7) mg/dL which decreased slightly to 35.7mg/dL (24.4 – 64.3). Median NEO level increased nearly 50% from 321.9 (135.6 – 661.4) nmol/L at baseline to 499.9 (106.0 – 1,041.0) nmol/L at follow-up. Similarly, median MPO level increased from 2,544 (1,459—4,276) ng/mL at baseline to 3,883 (1,842—12,590) ng/mL at follow-up. Nearly half of participants had elevated levels of AAT (45.4%) and MPO (47.9%) at both baseline and follow-up, while nearly two-thirds (67.2%) had elevated NEO at both baseline and follow-up. AAT and MPO were correlated moderately instantaneously at both timepoints (baseline: r=0.40, p<0.001 and follow-up: r=0.34, p<0.001) (Supplementary Table 1). There were no correlations in the same biomarker between timepoints, but baseline NEO was correlated with follow-up MPO (r=0.24, p<0.01). AAT and NEO exhibited circadian patterns (Supplementary Table 3) whereby stools taken during the daytime (6am – 6pm) had higher concentration of AAT (β = 0.67, 95% CI = 0.26 to 1.08. p<0.01) and lower NEO (β = −0.52, 95% CI = −0.96 to −0.08, p<0.05) compared to stools taken in the late evening or early morning. All multivariate analysis included adjustment for bowel motion time of day.

### Biomarker distributions across key covariates

The distribution of biomarker values across key covariates is presented in Figure 1A-N. AAT levels were not correlated with age and showed little variation in their distribution over time. NEO and MPO showed significant negative correlations with age but an upward shift to higher levels at follow-up across all ages, meaning the biomarker levels in 10–15-month-olds at follow-up were like infants 5-months younger at baseline. AAT and MPO levels were highly left-skewed, and NEO was moderately left-skewed. No significant differences in any biomarker level were observed between infants whose mother had or had not reported their infant having a fever in the last 3-months. Recent 7-day diarrhoea however was associated with elevated NEO at baseline (p<0.001) but not follow-up, and increased MPO at follow-up (p<0.001) but not baseline. Infants who were consuming high (≥ 1.5 serves) ASF showed lower NEO at follow-up (p<0.05) but no differences in AAT or MPO. Infants who were currently receiving breastmilk had raised mean biomarker values which were statistically significant for AAT at baseline (p<0.001), NEO at follow-up (p<0.05) and MPO at baseline (p<0.05) and follow-up (p<0.05). Breastfeeding status at baseline was able to predict higher levels of AAT, NEO, and MPO at follow-up independently of follow-up breastfeeding status indicating a temporal effect (i.e. not due to instantaneous contamination of stool samples with biomarkers from breastmilk) (Supplementary Table 5).

**Figure 1.**
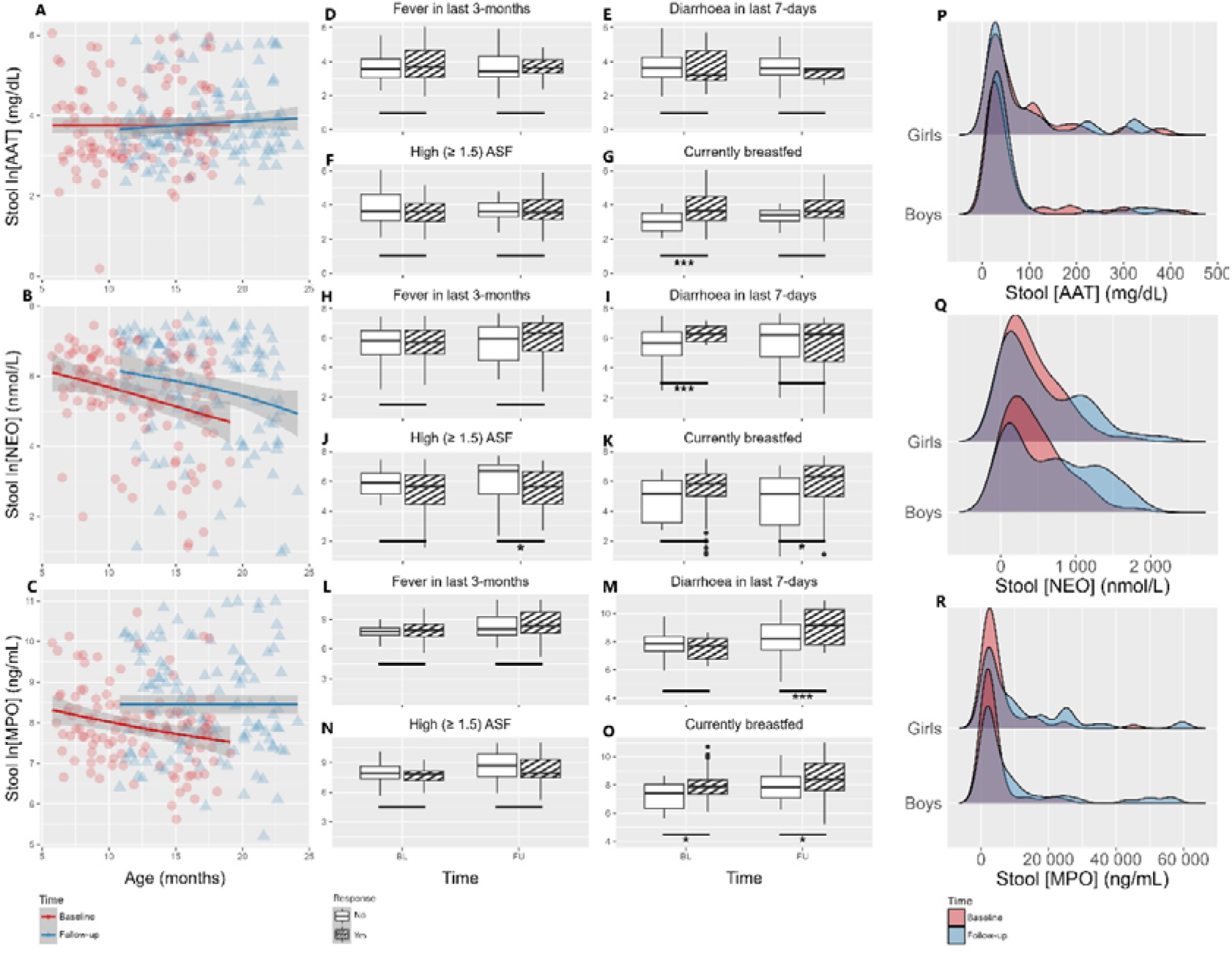
Distribution of EED biomarkers by age and sex, and key health covariates. **A-C:** Scatter plots of log-transformed biomarkers across age at baseline (red circles) and follow-up (blue triangles). Shaded grey line represents confidence interval for a simple linear model of biomarker value regressed against age. **D-O:** Boxplots of log-transformed biomarkers by key covariates – fever in last 3-months, diarrhoea in last 7-days, high animal-source food (ASF) intake, and current breastfeeding status, at baseline and follow-up. Shaded boxes represent the response a yes response. Asterisks represent p-values for a t-test comparing mean biomarker levels at each time point between the categories of the covariate response (no/yes). *p<0.05 **p<0.01 ***p<0.001.**P-R:** Kernel density plots of untransformed stool AAT, NEO, and MPO concentrations stratified by sex and time (red = baseline, blue = follow-up).

Other demographic and socio-economic variables were infrequently associated with biomarker concentrations. Mothers age in years was borderline associated with lower AAT (β = −0.02, 95% CI = −0.04 to 0.00, p=0.07) and lower MPO (β = 0.07, 95% CI = 0.02 to 0.12. p<0.01). Mothers’ education however was not associated with any of the biomarkers. Associations between other socio-economic variables did not conform to a clear pattern with high household expenses associated with higher AAT (β = 0.27, 95% CI = −0.01 to 0.54, p=0.06) and MPO (β = 0.41, 95% CI = −0.03 to 0.84, p=0.07). Infants whose household had a fridge had a drop in AAT (β_interaction_ = −0.37, 95% CI = −0.67 to −0.07, p<0.05) and drop in MPO (β_interaction_ = −0.55, 95% CI = −1.10 to −0.00, p<0.05). Notably, NEO concentrations were not significantly associated with any socio-economic variable.

### LMER modelling of EED biomarkers

Key household level core WASH variables were weakly associated with biomarker concentrations (Table 3-5). Unimproved sanitation was weakly associated with higher AAT (β = 0.21, 95% CI = −0.05 to 0.47, p=0.11). Partial dirt floors in the household were associated weakly with lower AAT (β = −0.29, 95% CI = −0.57 to −0.00, p<0.05) and strongly with lower MPO (β = −0.59, 95% CI = −0.91 to −0.27, p<0.001). Infants whose household were supplied with municipal water compared to spring water displayed a non-significant increase in AAT (β_interaction_ = 0.41, 95% CI = −0.10 to 0.92, p=0.11) resulting in borderline significantly higher AAT at follow-up (β = 0.36, 95% CI = −0.04, 0.76, p=0.08) and an increase in NEO (β_interaction_ = 0.67, 95% CI = −0.18 to 1.51, p=0.12) resulting in significantly higher NEO at follow-up (β = 0.71, 95% CI = 0.05, 1.37, p<0.05), and a significant increase in MPO over time (p_interaction_ < 0.05) resulting in higher MPO at follow-up (β = 0.56, 95% CI = 0.11 to 1.02, p<0.01). Unsafe disposal of infant faeces was not associated with AAT or MPO but was associated with a significant increase in NEO over time (p_interaction_ < 0.05) resulting in higher NEO at follow-up (β = 0.82, 95% CI = 0.20 to 1.43, p<0.01). Combined with the key covariates, demographic, and socio-economic variables, these core WASH variables models explained 25%, 26%, and 32% of the variance in AAT, NEO, and MPO respectively.

**Table 3.**
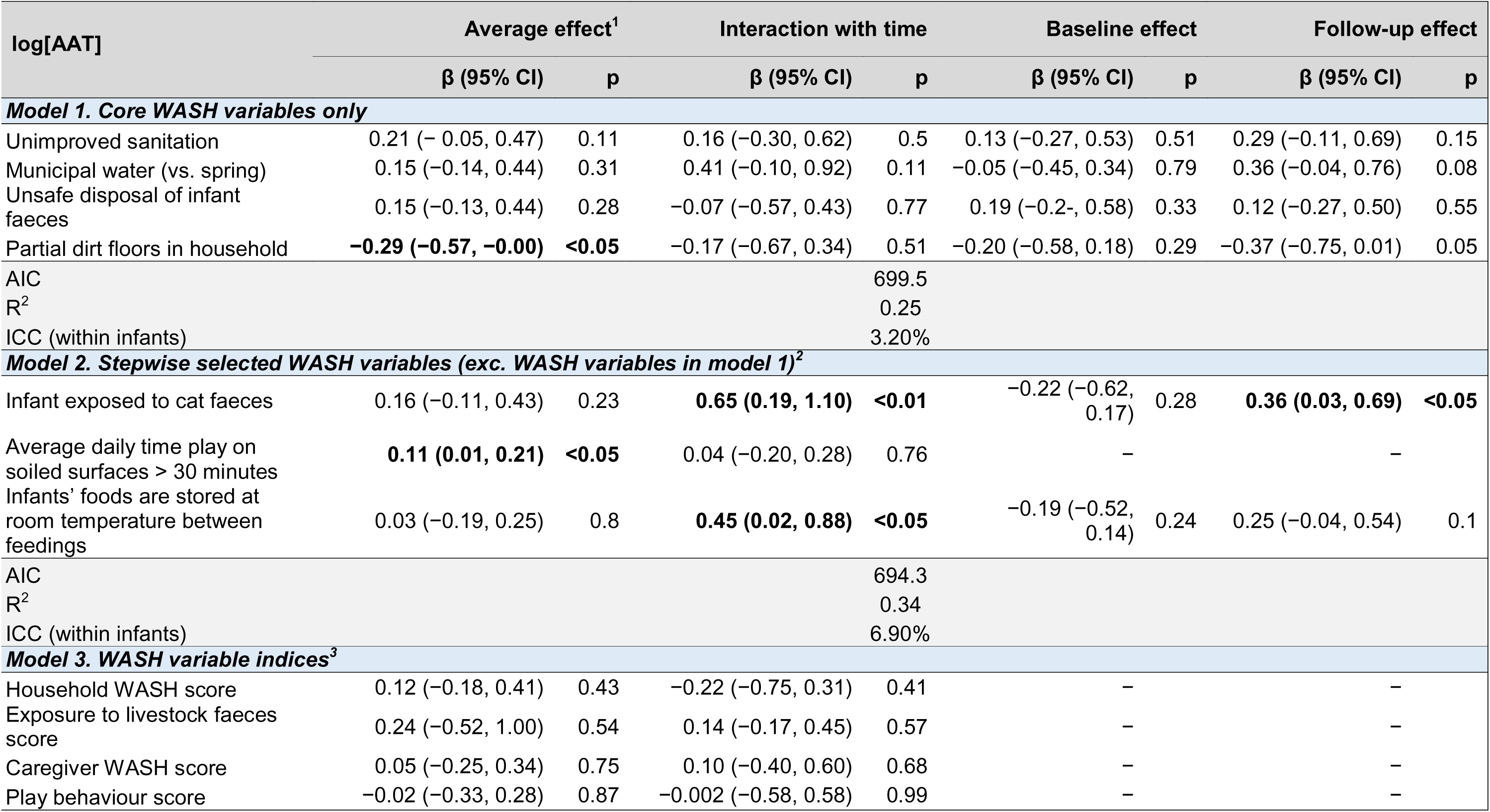
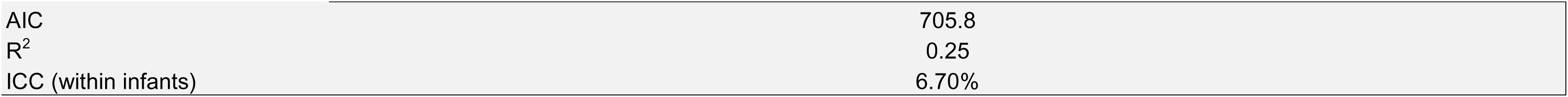
Linear mixed effects regression model of the associations between WASH variables and log-transformed AAT (mg/dL). All models are adjusted for age, sex, breastfeeding status, mothers age, animal source food intake, recent diarrhoea, fever, bowel motion time of day, and socio-economic variables. Baseline and follow-up isolated effects are modelled when the interaction with time is significant at p<0.10. Random intercepts are included at the infant and village level to take into account clustering and repeated measures. ^1^The average effect refers to the mean effect on AAT across both baseline and follow-up. ^2^Stepwise selected WASH variables do not include the core WASH variables and are selected from an array of variables less-established in the literature as posing a WASH-EED risk including: exposure to poultry, cat, bird faeces, open storage of infants foods at room temperature, average daily time infant plays on ground, mother washes hands before preparing meals, mouthing of soil/sand/dirt, mouthing of soiled fomites, and father handles livestock/livestock faeces. ^3^WASH variable indices are made up of the some of the binary variables constituting each WASH variable category where the high-risk outcome is coded as 1, reference coded as 0. A high score in these variables indicates greater risk to poor WASH. AIC, Akaike information criterion. ICC, inter-class correlation.

| log[AAT] | Average effect<sup>1</sup> |  | Interaction with time |  | Baseline effect |  | Follow-up effect |  |
| --- | --- | --- | --- | --- | --- | --- | --- | --- |
|  | β (95% CI) | p | β (95% CI) | p | β (95% CI) | p | β (95% CI) | p |
| **Model 1. Core WASH variables only** |  |  |  |  |  |  |  |  |
| Unimproved sanitation | 0.21 (− 0.05, 0.47) | 0.11 | 0.16 (−0.30, 0.62) | 0.5 | 0.13 (−0.27, 0.53) | 0.51 | 0.29 (−0.11, 0.69) | 0.15 |
| Municipal water (vs. spring) | 0.15 (−0.14, 0.44) | 0.31 | 0.41 (−0.10, 0.92) | 0.11 | −0.05 (−0.45, 0.34) | 0.79 | 0.36 (−0.04, 0.76) | 0.08 |
| Unsafe disposal of infant faeces | 0.15 (−0.13, 0.44) | 0.28 | −0.07 (−0.57, 0.43) | 0.77 | 0.19 (−0.2-, 0.58) | 0.33 | 0.12 (−0.27, 0.50) | 0.55 |
| Partial dirt floors in household | **−0.29 (−0.57, −0.00)** | **<0.05** | −0.17 (−0.67, 0.34) | 0.51 | −0.20 (−0.58, 0.18) | 0.29 | −0.37 (−0.75, 0.01) | 0.05 |
| AIC |  |  |  | 699.5 |  |  |  |  |
| R<sup>2</sup> |  |  |  | 0.25 |  |  |  |  |
| ICC (within infants) |  |  |  | 3.20% |  |  |  |  |
| **Model 2. Stepwise selected WASH variables (exc. WASH variables in model 1)<sup>2</sup>** |  |  |  |  |  |  |  |  |
| Infant exposed to cat faeces | 0.16 (−0.11, 0.43) | 0.23 | **0.65 (0.19, 1.10)** | **<0.01** | −0.22 (−0.62, 0.17) | 0.28 | **0.36 (0.03, 0.69)** | **<0.05** |
| Average daily time play on soiled surfaces > 30 minutes | **0.11 (0.01, 0.21)** | **<0.05** | 0.04 (−0.20, 0.28) | 0.76 | – |  | – |  |
| Infants' foods are stored at room temperature between feedings | 0.03 (−0.19, 0.25) | 0.8 | **0.45 (0.02, 0.88)** | **<0.05** | −0.19 (−0.52, 0.14) | 0.24 | 0.25 (−0.04, 0.54) | 0.1 |
| AIC |  |  |  | 694.3 |  |  |  |  |
| R<sup>2</sup> |  |  |  | 0.34 |  |  |  |  |
| ICC (within infants) |  |  |  | 6.90% |  |  |  |  |
| **Model 3. WASH variable indices<sup>3</sup>** |  |  |  |  |  |  |  |  |
| Household WASH score | 0.12 (−0.18, 0.41) | 0.43 | −0.22 (−0.75, 0.31) | 0.41 | – |  | – |  |
| Exposure to livestock faeces score | 0.24 (−0.52, 1.00) | 0.54 | 0.14 (−0.17, 0.45) | 0.57 | – |  | – |  |
| Caregiver WASH score | 0.05 (−0.25, 0.34) | 0.75 | 0.10 (−0.40, 0.60) | 0.68 | – |  | – |  |
| Play behaviour score | −0.02 (−0.33, 0.28) | 0.87 | −0.002 (−0.58, 0.58) | 0.99 | – |  | – |  |

**Table 3.** Linear mixed effects regression model of the associations between WASH variables and log-transformed **AAT** (mg/dL).
|  |  |
| --- | --- |
| AIC | 705.8 |
| R<sup>2</sup> | 0.25 |
| ICC (within infants) | 6.70% |

**Table 4.**
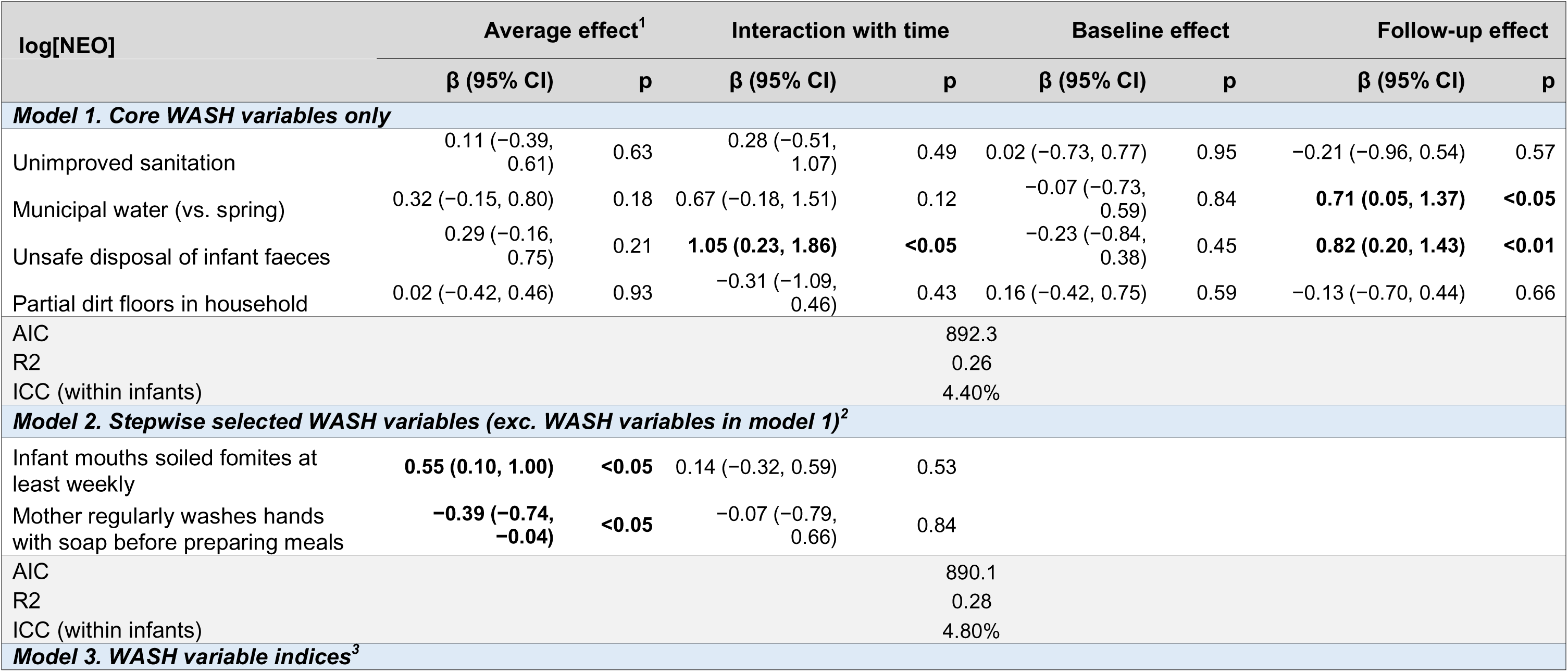
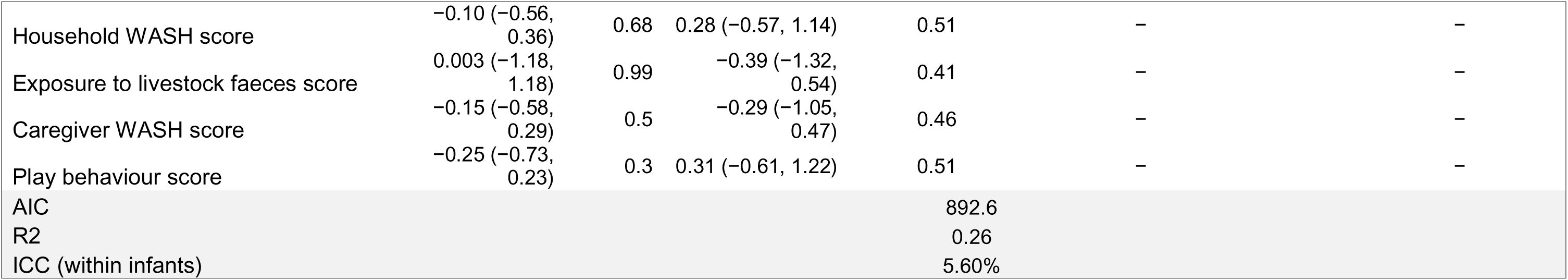
Linear mixed effects regression model of the associations between WASH variables and log-transformed **NEO** (nmol/L). All models are adjusted for age, sex, breastfeeding status, mothers age, animal source food intake, recent diarrhoea, fever, bowel motion time of day, and socio-economic variables. Baseline and follow-up isolated effects are modelled when the interaction with time is significant at p<0.10. Random intercepts are included at the infant and village level to take into account clustering and repeated measures. ^1^The average effect refers to the mean effect on NEO across both baseline and follow-up. ^2^Stepwise selected WASH variables do not include the core WASH variables and are selected from an array of variables less-established in the literature as posing a WASH-EED risk including: exposure to poultry, cat, bird faeces, open storage of infants foods at room temperature, average daily time infant plays on ground, mother washes hands before preparing meals, mouthing of soil/sand/dirt, mouthing of soiled fomites, and father handles livestock/livestock faeces. ^3^WASH variable indices are made up of the some of the binary variables constituting each WASH variable category where the high-risk outcome is coded as 1, reference coded as 0. A high score in these variables indicates greater risk to poor WASH. AIC, Akaike information criterion. ICC, inter-class correlation.

**Table 5.**
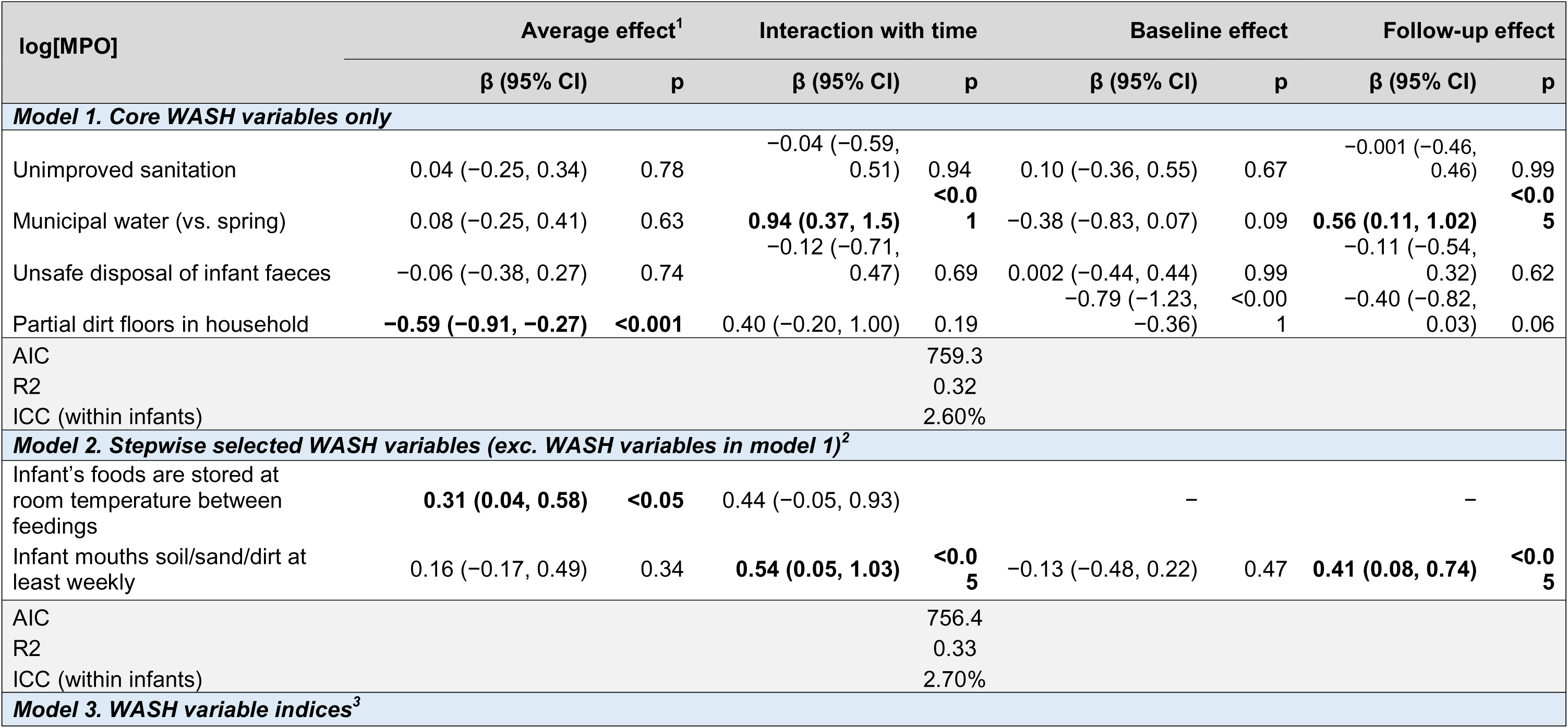
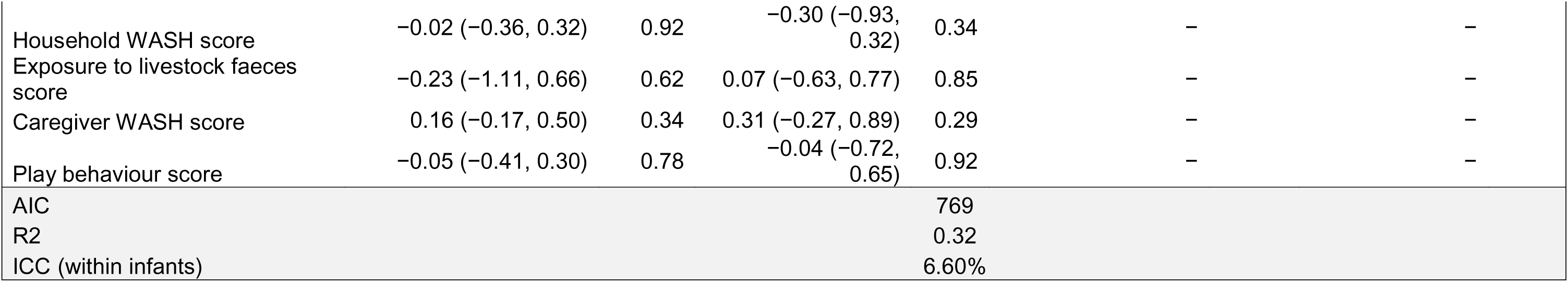
Linear mixed effects regression model of the associations between WASH variables and log-transformed **MPO** (ng/mL). All models are adjusted for age, sex, breastfeeding status, mothers age, animal source food intake, recent diarrhoea, fever, bowel motion time of day, and socio-economic variables. Baseline and follow-up isolated effects are modelled when the interaction with time is significant at p<0.10. Random intercepts are included at the infant and village level to take into account clustering and repeated measures. ^1^The average effect refers to the mean effect on MPO across both baseline and follow-up. ^2^Stepwise selected WASH variables do not include the core WASH variables and are selected from an array of variables less-established in the literature as posing a WASH-EED risk including: exposure to poultry, cat, bird faeces, open storage of infants foods at room temperature, average daily time infant plays on ground, mother washes hands before preparing meals, mouthing of soil/sand/dirt, mouthing of soiled fomites, and father handles livestock/livestock faeces. ^3^WASH variable indices are made up of the some of the binary variables constituting each WASH variable category where the high-risk outcome is coded as 1, reference coded as 0. A high score in these variables indicates greater risk to poor WASH. AIC, Akaike information criterion. ICC, inter-class correlation.

Hybrid stepwise selection of the exploratory WASH and play variables revealed some significant and consistent associations with EED biomarker levels. Infants whose complementary foods were regularly stored at room temperature between feedings always showed higher MPO (β = 0.31, 95% CI = 0.04 to 0.58, p<0.05) and an increase in AAT over time (β_interaction_ = 0.45, 95% CI = 0.02 to 0.88. p<0.05). Infants who play on soiled surfaces greater than 30 minutes daily had higher AAT (β = 0.11, 95% CI = 0.01, 0.21, p<0.05). Infants who mouthed soiled fomites at least weekly displayed significantly higher NEO (β = 0.55, 95% CI = 0.10 to 1.00, p<0.05) whilst infants who mouthed soil at least weekly displayed an increase in MPO over time (p_interaction_ < 0.05) resulting in higher MPO at follow-up (β = 0.41, 95% CI = 0.08 to 0.74, p<0.05). Exposure to cat faeces was associated with an increase in AAT over time (p_interaction_ < 0.01) leading to higher AAT at follow-up (β = 0.36, 95% CI = 0.03 to 0.69, p<0.05). and infants whose mothers regularly washed their hands with soap before preparing meals had lower NEO (β = −0.39, 95% CI = −0.74 to −0.04, p<0.05).

None of the WASH variable indices were associated with EED biomarker concentrations with all p-values of the average effects exceeding 0.30.

### Cross-sectional and temporal associations between EED and anthropometry

The adjusted association between anthropometry and EED biomarkers revealed mixed results (Tables 6-8).

**Table 6.** Linear mixed-effects regression (LMER) modelling of the cross-sectional and longitudinal association between anthropometry and log-transformed **AAT** (mg/dL). The LMER is an extension of the socio-economic and key covariate adjusted LMER used in the measurement of the association between WASH and EED with random intercepts for infant and village to account for clustering and repeated measurements. HAZ and WHZ are modelled jointly. ^1^The average effect refers to the mean association between the predictor and AAT across both time points. ^2^The instantaneous, or cross-sectional effect, is the association between infants’ attained anthropometry and biomarker level at the same point in time. ^3^The temporal effect considers the association between baseline anthropometry specifically on follow-up biomarker levels.

| Anthropometric indicators | Average effect<sup>1</sup> |  | Interaction with time |  | Baseline effect |  | Follow-up effect |  |
| --- | --- | --- | --- | --- | --- | --- | --- | --- |
| | $\beta$ (95% CI) | p | $\beta$ (95% CI) | p | $\beta$ (95% CI) | p | $\beta$ (95% CI) | p |
| **Model 1: Baseline and follow-up anthropometry (instantaneous effect) on $\ln[AAT]$<sup>2</sup>** |  |  |  |  |  |  |  |  |
| **Anthropometry z-scores stratified** |  |  |  |  |  |  |  |  |
| HAZ > -1 vs. -2 to -1 | -0.11 (-0.41, 0.19) | 0.49 | **0.59 (0.04, 1.14)** | **<0.05** | **-0.43 (-0.82, -0.05)** | **<0.05** | 0.16 (-0.29, 0.61) | 0.63 |
| HAZ ≤ -2 vs. -2 to -1 | -0.03 (-0.36, 0.29) | 0.83 | 0.05 (-0.58, 0.68) | 0.87 | -0.08 (-0.52, 0.35) | 0.71 | -0.03 (-0.50, 0.44) | 0.90 |
| WHZ > 0 (vs. ≤ 0) | 0.02 (-0.23, 0.28) | 0.85 | -0.27 (-0.75, 0.20) | 0.26 | 0.17 (-0.18, 0.52) | 0.34 | -0.10 (-0.44, 0.24) | 0.56 |
| **Continuous predictors** |  |  |  |  |  |  |  |  |
| HAZ | 0.05 (-0.09, 0.19) | 0.47 | 0.22 (-0.02, 0.46) | 0.07 | -0.05 (-0.23, 0.13) | 0.59 | 0.17 (-0.02, 0.36) | 0.08 |
| WHZ | 0.07 (-0.06, 0.20) | 0.28 | -0.16 (-0.40, 0.08) | 0.2 | 0.14 (-0.03, 0.32) | 0.10 | -0.01 (-0.19, 0.17) | 0.89 |
| **Model 2: Baseline anthropometry only (temporal effect) on follow-up $\ln[AAT]$<sup>3</sup>** |  |  |  |  |  |  |  |  |
| **Anthropometry z-scores stratified** |  |  |  |  |  |  |  |  |
| HAZ<sub>BL</sub> > -1 vs. -2 to -1 |  |  |  |  |  |  | 0.07 (-0.34, 0.48) | 0.74 |
| HAZ<sub>BL</sub> ≤ -2 vs. -2 to -1 | | | | | | | **-0.39 (-0.72, -0.05)** | **<0.05** |
| WHZ<sub>BL</sub> > 0 (vs. ≤ 0) | | | | | | | -0.25 (-0.60, 0.11) | 0.17 |
| **Continuous predictors** |  |  |  |  |  |  |  |  |
| HAZ<sub>BL</sub> |  |  |  |  |  |  | **0.17 (0.01, 0.32)** | **<0.05** |
| WHZ<sub>BL</sub> |  |  |  |  |  |  | -0.03 (-0.20, 0.15) | 0.76 |

**Table 7.** Linear mixed-effects regression (LMER) modelling of the cross-sectional and longitudinal association between anthropometry and log-transformed **NEO** (nmol/L). The LMER is an extension of the socio-economic and ke y covariate adjusted LMER used in the measurement of the association between WASH and EED with random intercepts for infant and village to account for clustering and repeated measurements. HAZ and WHZ are modelled jointly. ^1^The average effect refers to the mean association between the predictor and NEO across both time points. ^2^The instantaneous, or cross-sectional effect, is the association between infants’ attained anthropometry and biomarker level at the same point in time. ^3^The temporal effect considers the association between baseline anthropometry specifically on follow-up biomarker levels.

| Anthropometric indicators | Average effect<sup>1</sup> |  | Interaction with time |  | Baseline effect |  | Follow-up effect |  |
| --- | --- | --- | --- | --- | --- | --- | --- | --- |
| | $\beta$ (95% CI) | p | $\beta$ (95% CI) | p | $\beta$ (95% CI) | p | $\beta$ (95% CI) | p |
| **Model 1: Baseline and follow-up anthropometry (instantaneous effect) on $\ln[\text{NEO}]^2$** |  |  |  |  |  |  |  |  |
| **Anthropometry z-scores stratified** |  |  |  |  |  |  |  |  |
| HAZ > -1 vs. -2 to -1 | 0.14 (-0.32, 0.60) | 0.54 | 0.18 (-0.70, 1.06) | 0.69 | 0.04 (-0.63, 0.70) | 0.91 | 0.22 (-0.39, 0.83) | 0.48 |
| HAZ ≤ -2 vs. -2 to -1 | 0.01 (-0.49, 0.50) | 0.98 | -0.28 (-1.26, 0.70) | 0.58 | 0.12 (-0.56, 0.80) | 0.73 | -0.16 (-0.87, 0.56) | 0.66 |
| WHZ > 0 (vs. ≤ 0) | -0.22 (-0.61, 0.17) | 0.27 | -0.41 (-1.12, 0.34) | 0.28 | -0.01 (-0.55, 0.53) | 0.97 | -0.42 (-0.95, 0.11) | 0.12 |
| **Continuous predictors** |  |  |  |  |  |  |  |  |
| HAZ | 0.17 (-0.04, 0.39) | 0.11 | 0.07 (-0.32, 0.46) | 0.73 | 0.14 (-0.13, 0.42) | 0.31 | 0.21 (-0.09, 0.51) | 0.17 |
| WHZ | -0.07 (-0.27, 0.13) | 0.47 | -0.07 (-0.45, 0.32) | 0.72 | -0.04 (-0.31, 0.23) | 0.78 | -0.11 (-0.39, 0.17) | 0.45 |
| **Model 2: Baseline anthropometry only (temporal effect) on follow-up $\ln[\text{NEO}]^3$** |  |  |  |  |  |  |  |  |
| **Anthropometry z-scores stratified** |  |  |  |  |  |  |  |  |
| HAZ<sub>BL</sub> > -1 vs. -2 to -1 |  |  |  |  |  |  | 0.33 (-0.32, 0.97) | 0.32 |
| HAZ<sub>BL</sub> ≤ -2 vs. -2 to -1 | | | | | | | -0.41 (-1.09, 0.28) | 0.24 |
| WHZ<sub>BL</sub> > 0 (vs. ≤ 0) | | | | | | | -0.10 (-0.65, 0.46) | 0.74 |
| **Continuous predictors** |  |  |  |  |  |  |  |  |
| HAZ<sub>BL</sub> |  |  |  |  |  |  | **0.36 (0.08, 0.63)** | **<0.05** |
| WHZ<sub>BL</sub> |  |  |  |  |  |  | -0.19 (-0.46, 0.09) | 0.18 |

**Table 8.** Linear mixed-effects regression (LMER) modelling of the cross-sectional and longitudinal association between anthropometry and log-transformed **MPO** (mg/g). The LMER is an extension of the socio-economic and key covariate adjusted LMER used in the measurement of the association between WASH and EED with random intercepts for infant and village to account for clustering and repeated measurements. HAZ and WHZ are modelled jointly. ^1^The average effect refers to the mean association between the predictor and MPO across both time points. ^2^The instantaneous, or cross-sectional effect, is the association between infants’ attained anthropometry and biomarker level at the same point in time. ^3^The temporal effect considers the association between baseline anthropometry specifically on follow-up biomarker levels.

| Anthropometric indicators | Average effect<sup>1</sup> |  | Interaction with time |  | Baseline effect |  | Follow-up effect |  |
| --- | --- | --- | --- | --- | --- | --- | --- | --- |
| | $\beta$ (95% CI) | p | $\beta$ (95% CI) | p | $\beta$ (95% CI) | p | $\beta$ (95% CI) | p |
| **Model 1: Baseline and follow-up anthropometry (instantaneous effect) on $\ln[MPO]$<sup>2</sup>** |  |  |  |  |  |  |  |  |
| **Anthropometry z-scores stratified** |  |  |  |  |  |  |  |  |
| HAZ > -1 vs. -2 to -1 | 0.29 (-0.04, 0.63) | 0.08 | **0.68 (0.05, 1.31)** | **<0.05** | -0.09 (-0.65, 0.46) | 0.88 | **0.59 (0.08, 1.09)** | **<0.05** |
| HAZ ≤ -2 vs. -2 to -1 | **0.31 (0.01, 0.62)** | **<0.05** | 0.42 (-0.29, 1.13) | 0.25 | 0.09 (-0.41, 0.58) | 0.70 | **0.50 (0.02, 0.99)** | **<0.05** |
| WHZ > 0 (vs. ≤ 0) | -0.27 (-0.56, 0.01) | 0.06 | **-0.59 (-1.12, -0.05)** | **<0.05** | 0.05 (-0.36, 0.45) | 0.82 | **-0.54 (-0.93, -0.15)** | **<0.01** |
| **Continuous predictors** |  |  |  |  |  |  |  |  |
| HAZ | 0.06 (-0.10, 0.22) | 0.46 | 0.16 (-0.13, 0.45) | 0.28 | -0.01 (-0.22, 0.20) | 0.91 | 0.15 (-0.08, 0.37) | 0.20 |
| WHZ | -0.09 (-0.24, 0.06) | 0.22 | -0.17 (-0.46, 0.11) | 0.23 | -0.01 (-0.21, 0.19) | 0.93 | -0.18 (-0.39, 0.02) | 0.08 |
| **Model 2: Baseline anthropometry only (temporal effect) on follow-up $\ln[MPO]$<sup>3</sup>** |  |  |  |  |  |  |  |  |
| **Anthropometry z-scores stratified** |  |  |  |  |  |  |  |  |
| HAZ<sub>BL</sub> > -1 vs. -2 to -1 |  |  |  |  |  |  | 0.06 (-0.43, 0.55) | 0.81 |
| HAZ<sub>BL</sub> ≤ -2 vs. -2 to -1 | | | | | | | -0.06 (-0.57, 0.45) | 0.83 |
| WHZ<sub>BL</sub> > 0 (vs. ≤ 0) | | | | | | | **-0.42 (-0.84, -0.01)** | **<0.05** |
| **Continuous predictors** |  |  |  |  |  |  |  |  |
| HAZ<sub>BL</sub> |  |  |  |  |  |  | 0.11 (-0.10, 0.32) | 0.31 |
| WHZ<sub>BL</sub> |  |  |  |  |  |  | -0.13 (-0.33, 0.08) | 0.23 |

Compared to infants at risk of stunting, stunted infants had no difference in levels of AAT when measured instantaneously at either time point. However, stunted infants at baseline had lower AAT levels at follow-up (β = −0.39, 95% CI = −0.72 to −0.05, p<0.05). Infants with HAZ > −1 compared to infants at risk of stunting at baseline had lower AAT (β = −0.43, 95% CI = −0.82 to −0.05, p<0.05) and this effect was not significant at follow-up.

HAZ and WHZ were not significantly associated with NEO at the time of measurement, although a strong but insignificant association was seen in infants with WHZ > 0 at follow-up associated with lower MPO at follow-up (β = −0.42, 95% CI = −0.95 to 0.11, p=0.12). A temporal effect was seen where HAZ at baseline modelled as a continuous variable was positively associated with higher NEO at follow-up (β = 0.36, 95% CI = 0.08 to 0.63, p<0.05).

The association between HAZ and MPO demonstrated a non-linear (U-shaped) pattern. Compared to infants at risk of stunting, stunted infants had higher MPO on average (β = 0.31, 95% CI = 0.01 to 0.62, p < 0.05) which was heavily skewed towards higher MPO at follow-up (β = 0.50, 95% CI = 0.02 to 0.99, p < 0.05) compared to baseline (β = 0.09, 95% CI = −0.41 to 0.58, p = 0.70). On the other hand, infants with high HAZ at follow-up also had higher MPO at follow-up (β = 0.59, 95% CI = 0.08 to 1.09, p < 0.05), generating this U-shaped pattern at follow-up. Lower MPO at follow-up was also seen in infants with WHZ > 0 (β = −0.54, 95% CI = −0.93 to −0.15, p<0.01). The associations between follow-up HAZ and follow-up MPO were not driven by prior anthropometry as baseline HAZ showed no association with follow-up MPO (High HAZ: β = 0.06, 95% CI = −0.43 to 0.55, p=0.81 and stunted: β = −0.06, 95% CI = −0.57 to 0.45, p=0.83). For WHZ > 0 however, baseline WHZ > 0 strongly predicted lower MPO at follow-up (β = −0.42, 95% CI = −0.84 to −0.01, p<0.05).

### Association between biomarkers and growth faltering

Biomarkers were not associated with the change in HAZ over the follow-up period (ΔHAZ) either when measured as a continuous variable or as tertile groups at baseline or follow-up (Supplementary Table 7). Higher NEO concentration at follow-up was associated with ΔHAZ but not statistically significant (β = −0.04, 95% CI = −0.09 to 0.01, p = 0.09). When modelled as a comparison of percentile groups, compared to infants with follow-up NEO between the 25^th^ to 75^th^ percentiles, infants with concentrations below the 25^th^ percentile had a borderline significant association with ΔHAZ (β = 0.18, 95% CI = −0.01 to 0.36, p = 0.06). There was a significant interaction between follow-up NEO and HAZ_BL_ (β_interaction_ = −0.05, 95% CI = −0.09 to −0.01, p < 0.05) which was driving a negative association between follow-up NEO and ΔHAZ amongst infants with baseline HAZ > −1 (β = −0.10, 95% CI = −0.18 to −0.02, p<0.05) (Supplementary Figure 1).

## Discussion

This study demonstrated that infants in rural Central Java, Indonesia, a population with little to no prior EED research, are experiencing extremely high levels of intestinal inflammation and protein loss and confirms the hypothesis that in this setting biomarkers of EED are both considerably elevated and associated with WASH conditions and anthropometry. Seasonality was observed in intestinal inflammation biomarkers MPO and NEO where mean concentrations were higher at follow-up (wet season) despite the older age of infants at follow-up and negative correlation between MPO and NEO with age. This contributes to some existing evidence (30, 54) of the slight seasonality of EED biomarkers. Stool AAT and MPO levels at baseline and follow-up were similar to levels seen in infants 0-5 years old in Bangladesh (55) and India (56) as well as multiple sites of the MAL-ED study (54) although the 75^th^ percentile of MPO seen at follow-up (12,590 ng/mL) in our study was far higher than these studies. Median NEO levels were slightly lower, but were still considered elevated in all but 17% of infants at baseline and were 20-50 times higher than healthy and norovirus-infected children < 5 years old in urban Indonesia (57).

Baseline mean HAZ was already low (−1.26) and overall HAZ only declined by an average of −0.09. This may explain why biomarkers did not exhibit significant associations with ΔHAZ as little growth faltering over the follow-up occurred. That follow-up NEO was only negatively associated with growth faltering in infants whose HAZ at baseline was above −1 is suggesting that most infants were likely past an EED biomarker-sensitive growth faltering period and only those infants with little previous growth faltering were susceptible.

The associations between attained HAZ and EED biomarkers were complex and differed depending on whether HAZ was modelled continuously or in categories and non-linear relationships were more commonly found. Whilst it is recognised that increasing severity of faecal pathogen exposure is proportional to EED severity and subsequently the degree of growth faltering, biomarkers of EED may be subject to nuance in their interpretation (31). The pattern observed for NEO is difficult to interpret but may reflect differences in immune maturation. Higher baseline HAZ was associated with higher NEO at follow-up, a finding that contrasts with the expectation that greater intestinal inflammation would be associated with poorer growth. It may be possible however that infants with improved growth at baseline mounted a more mature cell-mediated response to faecal pathogens at or close to follow-up, consistent with reports from a Brazilian cohort where elevated NEO in the absence of marked intestinal inflammation was associated with improved growth (58). However, both positive and negative associations between stool NEO levels and linear growth have been observed in other studies (59–63), thus the interpretation of NEO in relation to linear growth may be significantly affected by context and immune development.

The pattern of association between HAZ and AAT was indicative of a distinct time-dependent cross-over in barrier phenotype. Compared to infants at risk of stunting, those with the highest HAZ exhibited lower AAT at baseline and an increase at follow-up, whilst infants stunted at baseline showed a marked decline in AAT over time, resulting in lower AAT at follow-up, representing a cross-over in the trajectories between groups. This pattern is consistent with ealier epthielial barrier disruption and increased protein loss among infants who were already stunted at baseline, with subsequent barrier restitution (reduced luminal-to-mucosal leak) during follow-up leading to declining AAT. A similar temporal cross-over pattern has been reported in Zimbabwe, where Humphrey and colleagues have shown that biomarkers consistent with enterocyte injury, microbial translocation and systemic innate immune activation (e.g., I-FABP, sCD14, CRP, AGP) were elevated earlier in life among infants with poor linear growth but fell to levels below those of non-stunted children by 18-months, implying dynamic trajectories of mucosal injury and repair rather than a static EED state (32). In our cohort, the reduction in AAT amongst stunted infants could be due to partial resolution of permeability and enteropathy across the observation window, although this did not coincide with lower MPO at follow-up, suggesting that barrier restitution can occur in the presence of ongoing mucosal innate activation, or that AAT and MPO care capturing distinct processes of permeability vs. luminal neutrophilic inflammation. While our data cannot determine the precise timing of injury or repair phases, the observed pattern aligns with a model in which early-life barrier dysfunction and EED contributes to growth faltering, followed by incomplete or heterogeneous recovery, and that our sampling captured infants, on average, mid-transition along this trajectory.

In contrast, the MPO and HAZ relationship differed from AAT, and was best described as U-shaped at follow-up, with higher MPO in both infants with the highest HAZ and those stunted at follow-up, compared with infants at risk of stunting. Notably, MPO at follow-up was not associated with baseline HAZ or ΔHAZ, supporting the interpretation that MPO may reflect a proximal, episodic mucosal inflammatory biomarker, consistent with acute neutrophil recruitment and degranulation in response to recent enteric exposures, rather than cumulative chronic enteropathy. This is strengthened by the elevated MPO among infants with lower WHZ (acute undernutrition) at follow-up, linking MPO more closely to recent illness and acute undernutrition than to longer-term linear growth trajectories. The biological basis of the U-shaped relationship is unclear. It may reflect recent pathogen exposure accompanying infant increased mobility and environmental contact, whereas elevated MPO among stunted infants may reflect ongoing inflammatory burden, recurrent infections or altered mucosal function after barrier injury. Some studies have identified that elements of EED may be a survival advantage in the form of mucosal “containment” adaptation in the context of chronic pathogen exposure amongst infants with particularly poor linear growth (64). Residual confounding may also be a possibility in the associations reported, including differential caregiving and exposure pattern where infants with better growth may subsequently have fewer restrictions on play (increasing exposure of contaminated surfaces), while caregivers may increase close-contact care and reduce environmental exposure for infants who have become stunted, a mechanism that could influence luminal neutrophilia (MPO) independently of chronic barrier permeability (AAT).

We observed both previously recognised and newly recognised WASH factors associated with EED biomarkers and some similarities in the ambiguity of association directions as seen with biomarkers and linear growth. Spring water source (vs. municipal water) was associated with higher MPO at baseline, but a decrease in MPO and a significant increase in NEO over the follow-up period (Table 3). Unsafe water source has been associated with increased intestinal permeability (65), but existing literature does not explain these shifts over time that we saw amongst our study population. It may be that exposure to pathogens in spring water leads to a varied immune response over time with increased MPO initially and a cell-mediated (NEO) immune response as infants aged. It is difficult to juxtapose spring water as ‘unimproved’ and municipal water as ‘improved’ in this setting as in Indonesia (66) and noted elsewhere (67) municipal piped water supplies are frequently contaminated, although the negative association between spring water source and HAZ (Supplementary Table 5) would suggest it is more hazardous. A novel finding from our study was the association between prolonged storage of complementary foods and elevated AAT and MPO; a risk factor that previously has been associated with diarrhoea (68) as well as elevated *E. coli* levels in complementary foods (69) but to our knowledge not EED. This practice is common in mothers with low hygiene education and/or who are pressed for time and must prepare foods in the morning despite not owning a fridge. Ownership of a fridge was associated with a reduction in AAT and MPO over the follow-up which could be related to the hygienic preparation of infants’ complementary foods (69). In addition to these factors, infants whose mothers washed their hands before preparing foods had lower NEO, although this may be confounded by general mothers’ hygiene knowledge and practice. Overall, these findings indicate a major role of complementary food hygiene in driving elevated biomarker levels. Our finding that the mouthing of soil was associated with elevated MPO and soiled fomites elevated NEO is consistent with an emergence of studies highlighting the associations between enteric disease and inflammation from mouthing soil and soiled objects in contaminated environments in LMICs (70–72). Increased play on dirt and soiled surfaces was associated with elevated NEO highlighting the large possibility play in these contaminated environments drives elevated intestinal inflammation.

Considering these positive associations between mouthing of soil, soiled fomites, and play time on soil/dirt surfaces, the finding that presence of dirt floors in the household was associated strongly with lower AAT and MPO and improved linear growth over the follow-up period (Supplementary Table 5) is surprising. Presence of dirt floors has been associated in other studies with diarrhoea (73) and stunting (74), suggesting that presence of dirt floors in this study may be confounded by unmeasured factors. It may be that households with partial dirt floors were in more remote corners of the village leading to reduced pathogen exposure, but this remains postulation.

The lack of association between unimproved household sanitation and EED biomarkers or linear growth is likely since unimproved latrines in this area were typically piped to surrounding fishponds or rivers and therefore not posing an immediate health risk to infants. Exposure to cat faeces, but not poultry faeces, was associated with elevated EED biomarkers which may be a function of frequency and intensity of exposure with stray cats more common than uncaged chickens and ducks in these villages.

An incidental finding of this analysis was that breastmilk consumption predicted higher AAT, NEO, and MPO concentrations. This may reflect residual confounding whereby mothers who discontinued breastfeeding differ in hygiene practices, complementary food provision, and daily pathogen exposure patterns. It is unlikely that microquantities of biomarkers present in breastmilk ‘contaminate’ stool samples as the concentrations are too small in comparison as also argued by McCormick and colleagues (54), which is also suggested by the finding that baseline breastfeeding status predicts higher follow-up biomarker levels independent of follow-up breastfeeding status (Supplementary Table 5). Rather, it is possible this is driven by genuine biological effects of breastmilk on mucosal immunity and microbial exposure, rather than a direct causal increase in enteropathy severity. Breastmilk can reshape the mucosal immune landscape and microbial ecology in ways that could shift biomarker readouts without necessarily worsening structural EED. Breastmilk delivers IgA, antimicrobial factors (e.g., lactoferrin, lysozyme), cytokines/growth factors, and human milk oligosaccharides, which collectively modulate microbial colonisation, epithelial barrier maturation, and immune education (75). These inputs can alter the balance between immune “containment” and immune activation, such as changes in microbial exposure patterns and antigen sampling may bias toward IFN-γ –linked myeloid activation (potentially increasing neopterin, a Th1-associated macrophage activation marker) even while improving certain barrier functions (76). Accordingly, we interpret breastfeeding associations cautiously as potentially reflecting residual confounding and/or immunological programming effects, and not necessarily a causal increase in enteropathy severity.

We found infants who had diarrhoea within the last week had significantly higher NEO at baseline and MPO at follow-up in their second non-diarrhoeal stool sample taken up to a week later (Supplementary Table 2). This is an interesting observation that future studies would also consider as studies that exclude infants with diarrhoea on the day of data collection and therefore don’t include recent diarrhoea in their analysis (77) would fail to adjust for this significant contributor.

The high levels of intestinal inflammation and permeability seen amongst infants in this study is quite striking considering the overall uniform coverage of at least unimproved sanitation in these villages and at least piped water available in every household. In this area of Indonesia, the main stunting programs that are funded and implemented by local health centres and villages are related to complementary food supplementation and vaccination but not WASH practices and infant hygiene (author observation). Overall, there is a serious systematic neglect towards ‘baby WASH’ and its contribution to sub-optimal child growth (16). A mother and infant centred ‘baby WASH’ intervention program is needed to address the likely chronic exposure to faecal pathogens that is occurring in these villages despite overall high sanitation coverage.

The findings of our research should be interpreted carefully considering the small sample size. Linear growth of children is affected by a multitude of environmental, nutrition, infection, and genetic factors and to explain small deviations in linear growth with stool biomarkers in a small sample size may lead to the loss of statistical significance in small regression coefficients. The three biomarkers here represent important domains but not all domains of EED, and so readers should not assume that the findings presented here represent associations between WASH and growth with EED holistically. The variables measured in this dataset explained only ∼30% of the variation in biomarker concentrations highlighting the possibility of other unmeasured WASH and non-WASH factors influencing their concentration. Infants had a relatively low HAZ at baseline, therefore there may exist other factors such as poor infant and young child feeding practices or infections that had played a role in growth faltering amongst participants prior to their recruitment into this study. A larger cohort study recruiting infants prior to birth of a larger sample size is therefore warranted to better understand the complex associations presented here. As discussed, infants who are or become stunted likely share *patterns* of EED biomarkers over time rather than similar levels at a given time point. Overall, a holistic evaluation of the multitude of potential pathways leading to exposure of gastrointestinal pathogens that may trigger EED and growth faltering and related consequences amongst rural infants is needed as existing WASH infrastructure coupled with stagnant progress is evidently insufficient.

## Data Availability

Data are available upon reasonable request to the authors

## Acknowledgements

We thank the study participants for giving up their time for the participation in this research project. We also thank local village heads, midwives and health cadres for their assistance in the project.

## CRediT statement

**Callum Lowe:** Conceptualization; data curation, formal analysis, investigation, methodology, project administration, writing – original draft, writing – review & editing. **Tony Arjuna:** methodology, writing – review & editing. **Mubasyir Hasanbasri**: methodology, writing – review & editing. **Haribondhu Sarma:** methodology, writing – review & editing. **I Nyoman Sutarsa:** writing – review & editing. **Severine Navarro:** writing – review & editing. **Darren Gray:** funding acquisition, methodology, writing – review & editing. **Matthew Kelly:** methodology, writing – review & editing.

## Funding

This work was funded by QIMR Berghofer Medical Research Institute

## Conflicts of interest

The authors have no conflicts of interest to declare.

## Notes

### Competing Interest Statement

The authors have declared no competing interest.

### Funding Statement

This study was funded by QIMR Berghofer Medical Research Institute.

### Author Declarations

This project received ethical approval from the Indonesian National Research and Innovation Agency (BRIN) Health Research Ethics Committee (Protocol #26032024004917) and the Australian National University Research Ethics Committee (Protocol H/2023/1123).

