## Supplementary Material for "Elevated levels of environmental enteric dysfunction biomarkers among rural Indonesian infants: associations with water, sanitation, hygiene and linear growth"

|  | **AAT_BL_** | **NEO_BL_** | **MPO_BL_** | **AAT_FU_** | **NEO_FU_** | **MPO_FU_** |
| --- | --- | --- | --- | --- | --- | --- |
| **AAT_BL_** | 1 |  |  |  |  |  |
| **NEO_BL_** | −0.01 | 1 |  |  |  |  |
| **MPO_BL_** | **0.40***** | 0.16 | 1 |  |  |  |
| **AAT_FU_** | 0.16 | 0.05 | 0.12 | 1 |  |  |
| **NEO_FU_** | −0.04 | 0.14 | 0.15 | 0.03 | 1 |  |
| **MPO_FU_** | 0.08 | **0.24**** | 0.09 | **0.34***** | 0.03 | 1 |

**Supplementary Table 1.** Pearson correlation between and within baseline and follow−up biomarkers. * p<0.05 ** p<0.01 *** p<0.001

|  | **ln[AAT] mg/dL** | | | | **ln[NEO] nmol/L** | | | | **ln[MPO] ng/mL** | | | |
| --- | --- | --- | --- | --- | --- | --- | --- | --- | --- | --- | --- | --- |
|  | **Average effect** | | **Interaction with time** | | **Average effect** | | **Interaction with time** | | **Average effect** | | **Interaction with time** | |
|  | **B (95% CI)** | **p** | **B (95% CI)** | **p** | **B (95% CI)** | **p** | **B (95% CI)** | **p** | **B (95% CI)** | **p** | **B (95% CI)** | **p** |
| **Null model** | **n/a** |  | **n/a** |  | **n/a** |  | **n/a** |  | **n/a** |  | **n/a** |  |
| ***Model fit*** |  |  |  |  |  |  |  |  |  |  |  |  |
| R-squared | 0/0.165 | | | | 0/0.138 | | | | 0/0.026 | | | |
| ICC (individual | 15.60% | | | | 12.80% | | | | 2.60% | | | |
| ICC (village) | 0.10% | | | | 0.10% | | | | 0 | | | |
| AIC | 670.4 | | | | 882.9 | | | | 760.3 | | | |
| **Base model** |  |  |  |  |  |  |  |  |  |  |  |  |
| Age | **0.04 (0.02, 0.08)** | **<0.001** | ns |  | **-0.07 (-0.13, -0.01)** | **<0.05** | ns |  | -0.01 (-0.05, 0.03) | 0.66 | ns |  |
| Sex | **0.30 (0.05, 0.55)** | **<0.05** | ns |  | -0.17 (-0.54, 0.20) | 0.35 | ns |  | **0.43 (0.13, 0.72)** | **<0.01** | ns |  |
| Currently breastfed | **0.51 (0.13, 0.89)** | **<0.01** | ns |  | **0.94 (0.38, 1.50)** | **<0.01** | ns |  | **0.55 (0.12, 0.98)** | **<0.05** | ns |  |
| ASF intake (serves per day) | -0.10 (-0.22, 0.02) | 0.10 | ns |  | -0.16 (-0.34, 0.02) | 0.07 | ns |  | **-0.17 (-0.30, -0.03)** | **<0.05** | ns |  |
| Mother's age | -0.02 (-0.04, 0.00) | 0.07 | ns |  | -0.003 (-0.04, 0.03) | 0.86 | ns |  | **-0.04 (-0.07, -0.01)** | **<0.05** | **0.07 (0.02, 0.12)** | **<0.01** |
| 7-day diarrhoea | -0.13 (-0.49, 0.24) | 0.50 | ns |  | **1.04 (0.21, 1.86)** | **<0.05** | **-1.39 (-2.5, -0.26)** | **<0.05** | -0.25 (-0.87, 0.38) | 0.43 | **1.03 (0.19, 1.88)** | **<0.01** |
| Daytime stool (vs. nighttime) | **0.67 (0.26, 1.08)** | **<0.01** | **-0.78 (-1.36, -0.20)** | **<0.01** | **-0.52 (-0.96, -0.08)** | **<0.05** | ns |  | 0.01 (-0.33, 0.35) | 0.96 | ns |  |
| Fever in last 3-months | 0.19 (-0.05, 0.43) | 0.12 | ns |  | 0.14 (-0.23, 0.51) | 0.46 | ns |  | 0.18 (-0.10, 0.46) | 0.20 | ns |  |
| ***Model fit*** |  |  |  |  |  |  |  |  |  |  |  |  |
| R-squared | 0.14/0.21 | | | | 0.17/0.17 | | | | 0.20/0.23 | | | |
| ICC (individual | 8.70% | | | | 0% | | | | 4.70% | | | |
| ICC (village) | 0.40% | | | | 0.50% | | | | 0% | | | |
| AIC | 686.6 | | | | 881.6 | | | | 760.6 | | | |
| **SES-adjusted model** |  |  |  |  |  |  |  |  |  |  |  |  |
| ***Core variables*** |  |  |  |  |  |  |  |  |  |  |  |  |
| Mother has high education^1^ | -0.13 (-0.41, 0.16) | 0.38 | ns |  | 0.10 (-0.50, 0.70) | 0.74 | ns |  | 0.08 (-0.25, 0.41) | 0.64 | ns |  |
| High household expenses^2^ | 0.27 (-0.01, 0.54) | 0.06 | ns |  | 0.23 (-0.19, 0.65) | 0.28 | ns |  | 0.41 (-0.03, 0.84) | 0.07 | **-1.00 (-1.58, -0.42)** | **<0.001** |
| Father's job^3^ | -0.28 (-0.59, 0.02) | 0.07 | ns |  | 0.06 (-0.43, 0.55) | 0.81 | ns |  | -0.07 (-0.55, 0.40) | 0.76 | **0.75 (0.12, 1.38)** | **<0.05** |
| ***Other SES indicators*** |  |  |  |  |  |  |  |  |  |  |  |  |
| High bedroom count | ns |  |  |  | ns |  |  |  | ns |  |  |  |
| Own livestock | ns |  |  |  | ns |  |  |  | ns |  |  |  |
| Finished walls | ns |  |  |  | ns |  |  |  | ns |  |  |  |
| Own Wi-Fi | **0.30 (0.04, 0.56)** | **<0.05** | ns |  | ns |  |  |  | ns |  |  |  |
| Own water heater | ns |  |  |  | ns |  |  |  | ns |  |  |  |
| Own fridge | 0.26 (-0.09, 0.61) | 0.15 | **-0.37 (-0.67, -0.07)** | **<0.05** | ns |  |  |  | 0.04 (-0.37, 0.45) | 0.84 | **-0.55 (-1.10, -0.00)** | **<0.05** |
| ***Model fit*** |  |  |  |  |  |  |  |  |  |  |  |  |
| R-squared | 0.25 | | | | 0.25 | | | | 0.31 | | | |
| ICC (individual | 6.20% | | | | 4.80% | | | | 6.60% | | | |
| ICC (village) | 0 | | | | 0.27% | | | | 0 | | | |
| AIC | 693.0 | | | | 882.8 | | | | 757.6 | | | |

**Supplementary table 2.** Linear mixed effects regression modelling of the effects of demographic, socio-economic status (SES), and key covariates on log-transformed EED biomarker concentrations. All models include a random intercept for infant and for village in order to take into account clustering and repeat biomarker measures. R-squared values are written as the marginal/conditional variance explained. ICC; intra-class correlation at the individual and village level. AIC; Akaike information criterion. The SES-adjusted model includes all base model covariates. ns; not significant. Average effect refers to the mean effect of the covariate on baseline and follow-up biomarker levels in the pooled dataset.

^1^High education defined as Secondary or higher

^2^High household expenses defined as > Rp. 3.000.000 per month

^3^Fathers job where the exposure is employee or government employee and the reference is farmer, manual labour, or other occupation.

|  | **HAZ_BL_** | |  | **∆HAZ** | |
| --- | --- | --- | --- | --- | --- |
| **Covariates** | β (SE) | p |  | β (SE) | p |
| Age (months) | −0.02 (0.02) | 0.43 |  | −0.01 (0.02) | 0.39 |
| Female sex | **0.57 (0.16)** | **<0.001** |  | 0.01 (0.09) | 0.94 |
| Currently breastfed | −0.56 (0.29) | 0.05 |  | −0.27 (0.14) | 0.06 |
| Infant is mothers first born child | 0..23 (0.18) | 0.21 |  | −0.05 (0.12) | 0.67 |
| Father in higher earning occupation | 0.37 (0.23) | 0.11 |  | **−0.23 (0.10)** | **<0.05** |
| Mother has high education | −0.06 (0.19) | 0.76 |  | −0.14 (0.10) | 0.17 |
| Father has high education | **−0.39 (0.19)** | **<0.05** |  | 0.04 (0.10) | 0.69 |
| Household has high expenses | −0.28 (0.17) | 0.10 |  | **−0.28 (0.09)** | **<0.01** |
| Household assets – Wi-Fi | −0.25 (0.18) | 0.16 |  | **−0.20 (0.09)** | **<0.05** |
| Household owns livestock | 0.07 (0.17) | 0.67 |  | 0.05 (0.09) | 0.57 |
| Mother’s age (years) | −0.003 (0.02) | 0.86 |  | −0.002 (0.01) | 0.81 |
| House has finished walls | 0.08 (0.21) | 0.69 |  | −0.08 (0.11) | 0.46 |
| Household has high (≥3) bedroom count | 0.32 (0.18) | 0.07 |  | **0.19 (0.09)** | **<0.05** |
| Household assets — water heater | 0.43 (0.22) | 0.05 |  | **0.28 (0.12)** | **<0.05** |
| Mother’s height (cm) | **0.05 (0.02)** | **<0.001** |  | **0.02 (0.01)** | **<0.05** |
| Birthweight (g) | **0.62 (0.19)** | **<0.01** |  | −0.01 (0.10) | 0.92 |

**Supplementary Table 3.** Effect of age, sex, demographic and socio-economic factors on HAZ at baseline and ∆HAZ. Model is a simple linear regression for both outcomes, and all covariates are mutually adjusted. ∆HAZ model adjusts for HAZ_BL_.

|  | **Effect on HAZ_BL_** | | | |  | **Effect on ΔHAZ** | | | |
| --- | --- | --- | --- | --- | --- | --- | --- | --- | --- |
| **WASH variables** | Partially adjusted | | Fully adjusted | |  | Partially adjusted | | Fully adjusted | |
|  | β (95% CI) | p | β (95% CI) | p |  | β (95% CI) | p | β (95% CI) | p |
| **Household WASH conditions** |  |  |  |  |  |  |  |  |  |
| Spring water source (vs. municipal) | −0.22 (−0.57, 0.13) | 0.20 | **−0.27 (−0.52, −0.02)** | **<0.05** |  | −0.004 (−0.18, 0.17) | 0.97 | − |  |
| Unimproved sanitation | 0.12 (−0.19, 0.43) | 0.47 | − |  |  | −0.07 (−0.23, 0.09) | 0.39 | − |  |
| Infant faeces disposed unsafely | 0.01 (−0.34, 0.36) | 0.96 | − |  |  | −0.14 (−0.32, 0.04) | 0.12 | −0.11 (−0.27, 0.05) | 0.22 |
| Partial dirt/earth floors (vs. fully cemented/tiled) | −0.07 (−0.42, 0.28) | 0.72 | − |  |  | **0.18 (0.00, 0.36)** | **<0.05** | **0.18 (0.003, 0.36)** | **<0.05** |
| Father handles livestock or livestock faeces | 0.10 (−0.21, 0.41) | 0.52 | − |  |  | −0.11 (−0.27, 0.05) | 0.19 | −0.08 (−0.24, 0.08) | 0.33 |
| **Exposure to livestock** |  |  |  |  |  |  |  |  |  |
| Exposed to poultry faeces | 0.05 (−0.52, 0.62) | 0.86 | − |  |  | 0.09 (−0.16, 0.34) | 0.47 | − |  |
| Exposed to bird faeces | 0.44 (−0.01, 0.89) | 0.06 | 0.40 (−0.05, 0.85) | 0.09 |  | **0.22 (0.02, 0.42)** | **<0.05** | 0.05 (−0.17, 0.27) | 0.63 |
| Exposed to cat faeces | 0.09 (−0.26, 0.44) | 0.62 | **−** |  |  | **0.24 (0.08, 0.40)** | **<0.01** | **0.22 (0.04, 0.40)** | **<0.05** |
| **Mother’s WASH** |  |  |  |  |  |  |  |  |  |
| Mother washes hands with soap before preparing meals | −0.05 (−0.38, 0.28) | 0.76 | **−** |  |  | 0.03 (−0.15, 0.21) | 0.73 | − |  |
| Infants’ foods are stored at room temperature for prolonged periods | 0.15 (−0.16, 0.46) | 0.36 | − |  |  | 0.05 (−0.11, 0.21) | 0.53 | − |  |
| **Play behaviour** |  |  |  |  |  |  |  |  |  |
| Mouth soil/sand/dirt at least weekly | 0.22 (−0.33, 0.77) | 0.44 | **−** |  |  | −0.10 (−0.28, 0.08) | 0.25 | **−0.15 (−0.29, −0.01)** | **<0.05** |
| Mouth soiled fomites at least weekly | 0.23 (−0.16, 0.62) | 0.25 | 0.32 (−0.11, 0.75) | 0.15 |  | −0.03 (−0.21, 0.15) | 0.75 | − |  |
| Ever mouth animal faeces (vs. never) | 0.26 (−0.50, 1.02) | 0.50 | **−** |  |  | −0.01 (−0.46, 0.44) | 0.95 | − |  |
| Average daily play time on soiled surfaces > 30minutes^1^ | −0.05 (−0.44, 0.34) | 0.79 | **−** |  |  | **−0.21 (−0.41, −0.01)** | **<0.05** | −0.16 (−0.36, 0.04) | 0.12 |
| Infant plays on nearby roads | −0.31 (−0.70, 0.08) | 0.12 | **−0.31 (−0.60, −0.02)** | **<0.05** |  | −0.10 (−0.30, 0.10) | 0.33 | − |  |
| Infant plays in nearby gardens | 0.003 (−0.41, 0.41) | 0.99 | **−** |  |  | −0.02 (−0.24, 0.20) | 0.87 | − |  |

**Supplementary Table 4.** Effect of health and WASH predictors on attained height (HAZ) at baseline and ΔHAZ over the follow−up period modelled with linear regression. Partially adjusted model for HAZ_BL_ includes significant covariates in supplementary table 4. Fully adjusted model for HAZ_BL_ includes mutual adjustment for significant predictors at p<0.25 in the partially adjusted model. ΔHAZ partially adjusted model includes significant covariates in supplementary table 4. Fully adjusted ΔHAZ model includes these covariates and mutual adjustment for significant predictors at p<0.25 in the partially adjusted model. ASF; animal−source foods. SE; standard error.

|  | ln[AAT] mg/dL | |  | ln[NEO] nmol/L | |  | Ln[MPO] ng/mL | |
| --- | --- | --- | --- | --- | --- | --- | --- | --- |
| **Currently breastfed** | B (SE) | p |  | B (SE) | p |  | B (SE) | p |
| Baseline | 0.74 (−0.09, 1.54) | 0.07 |  | 1.20 (−0.19, 2.59) | 0.09 |  | 0.75 (−0.39, 1.89) | 0.20 |
| Follow-up | 0.05 (−0.62, 0.72) | 0.88 |  | 0.40 (−0.74, 1.54) | 0.49 |  | 0.18 (−0.76, 1.12) | 0.70 |

**Supplementary Table 5.** Impact of baseline breastfeeding status on follow-up biomarker levels adjusted for follow-up breastfeeding status. Model is an ordinary least squares linear regression adjusted for age and sex.

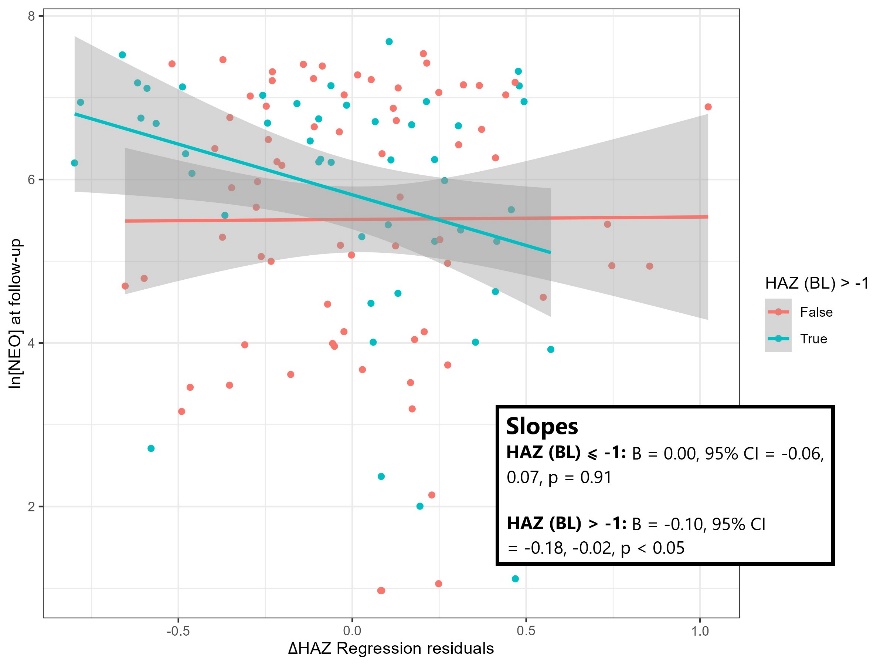

**Supplementary Figure 1**. Interaction between NEO levels at follow−up with HAZ at baseline on linear growth faltering (ΔHAZ) over the follow−up period. X−axis is the ΔHAZ linear regression residuals with NEO and the interaction term omitted.

|  | **[AAT] mg/dL** | | **[NEO] nmol/L** | | **[MPO] ng/mL** | |
| --- | --- | --- | --- | --- | --- | --- |
|  | **Baseline** | **Follow-up** | **Baseline** | **Follow-up** | **Baseline** | **Follow-up** |
| **Median (IQR)** | 37.2 (21.4—70.7) | 35.7 (24.4—64.3) | 321.9 (135.6—661.4) | 499.9 (106.0—1,041) | 2,544 (1,459—4,276) | 3,883 (1,842—12,590) |
| **Elevated concentrations^1^ N (%)** | 75 (63.0) | 81 (68.7) | 99 (83.2) | 94 (79.0) | 79 (66.4) | 85 (71.4) |
| **Elevated both time points** | 54 (45.4) | | 80 (67.2) | | 57 (47.9) | |

**Supplementary Table 6**. Median and interquartile ranges of untransformed biomarker values at baseline and follow-up and proportion of observations elevated in comparison to reference levels.

^1^Reference concentrations derived from McCormick et al (2017) as follows: AAT (<27 mg/dL), NEO (<70nmol/L), MPO (<2,000 ng/mL)

|  |  | | | | **Interaction with HAZ_BL_** | | | |
| --- | --- | --- | --- | --- | --- | --- | --- | --- |
|  | **Baseline** | | **Follow-up** | | **Baseline** | | **Follow-up** | |
|  | **β (95% CI)** | **p** | **β (95% CI)** | **p** | **β (95% CI)** | **p** | **β (95% CI)** | **p** |
| **ln[AAT] mg/dL** |  |  |  |  |  |  |  |  |
| **Continuous** | 0.01 (−0.07, 0.09) | 0.76 | 0.02 (−0.07, 0.11) | 0.68 | 0.06 (−0.02, 0.15) | 0.12 | 0.03 (−0.11, 0.17) | 0.69 |
| **Categorical** |  |  |  |  |  |  |  |  |
| **>75^th^** | −0.03 (−0.23, 0.17) | 0.79 | −0.08 (−0.27, 0.12) | 0.45 | − |  | − |  |
| **<25^th^** | −0.02 (−0.21, 0.18) | 0.87 | −0.09 (−0.30, 0.10) | 0.34 | − |  | − |  |
| **ln[NEO] nmol/L** |  | | | | | | | |
| **Continuous** | −0.01 (−0.07, 0.05) | 0.69 | −0.04 (−0.09, 0.01) | 0.09 | 0.01 (−0.07, 0.08) | 0.78 | **−0.05 (−0.09, −0.01)** | **<0.05** |
| **Categorical** |  |  |  |  |  |  |  |  |
| **>75^th^** | −0.06 (−0.26, 0.13) | 0.52 | 0.09 (−0.10, 0.29) | 0.35 | − |  | − |  |
| **<25^th^** | 0.07 (−0.15, 0.28) | 0.54 | 0.18 (−0.01, 0.38) | 0.07 | − |  | − |  |
| **ln[MPO] ng/mL** |  | | | | | | | |
| **Continuous** | 0.04 (−0.06, 0.13) | 0.45 | 0.02 (−0.04, 0.08) | 0.51 | −0.01 (−0.11, 0.10) | 0.9 | 0.01 (−0.05, 0.07) | 0.83 |
| **Categorical** |  |  |  |  |  |  |  |  |
| **>75^th^** | −0.07 (−0.27, 0.13) | 0.51 | 0.16 (−0.04, 0.35) | 0.12 | − |  | − |  |
| **<25^th^** | 0.02 (−0.18, 0.22) | 0.85 | 0.05 (−0.14, 0.25) | 0.6 | − |  | − |  |

**Supplementary Table 7**. Linear regression model of the association between EED biomarkers and change in growth (ΔHAZ) over the follow−up period. Model is adjusted for baseline HAZ, age, sex, breastfeeding status, mother’s height, mother’s education, household expenses, diarrhoea, fever, WASH, and animal source food intake. Each biomarker is modelled in mutual adjustment with other biomarkers, but a separate model is fitted for each time point.

|  | Item No | Recommendation | Page # |
| --- | --- | --- | --- |
| **Title and abstract** | 1 | (*a*) Indicate the study’s design with a commonly used term in the title or the abstract | 1 |
|  |  | (*b*) Provide in the abstract an informative and balanced summary of what was done and what was found | 2 |
| Introduction | | |  |
| Background/rationale | 2 | Explain the scientific background and rationale for the investigation being reported | 3 |
| Objectives | 3 | State specific objectives, including any prespecified hypotheses | 4-5 |
| Methods | | |  |
| Study design | 4 | Present key elements of study design early in the paper | 6 |
| Setting | 5 | Describe the setting, locations, and relevant dates, including periods of recruitment, exposure, follow-up, and data collection | 6-7 |
| Participants | 6 | (*a*) Give the eligibility criteria, and the sources and methods of selection of participants | 6-7 |
| Variables | 7 | Clearly define all outcomes, exposures, predictors, potential confounders, and effect modifiers. Give diagnostic criteria, if applicable | 7-8 |
| Data sources/ measurement | 8* | For each variable of interest, give sources of data and details of methods of assessment (measurement). Describe comparability of assessment methods if there is more than one group | *7-8* |
| Bias | 9 | Describe any efforts to address potential sources of bias | n/a |
| Study size | 10 | Explain how the study size was arrived at | 6 |
| Quantitative variables | 11 | Explain how quantitative variables were handled in the analyses. If applicable, describe which groupings were chosen and why | 8-9 |
| Statistical methods | 12 | (*a*) Describe all statistical methods, including those used to control for confounding | 8-11 |
|  |  | (*b*) Describe any methods used to examine subgroups and interactions | 8-11 |
|  |  | (*c*) Explain how missing data were addressed | n/a |
|  |  | (*d*) If applicable, describe analytical methods taking account of sampling strategy | 8-11 |
|  |  | (*e*) Describe any sensitivity analyses | 11 |
| Results | | |  |
| Participants | 13* | (a) Report numbers of individuals at each stage of study—eg numbers potentially eligible, examined for eligibility, confirmed eligible, included in the study, completing follow-up, and analysed | 12 |
|  |  | (b) Give reasons for non-participation at each stage | n/a |
|  |  | (c) Consider use of a flow diagram | n/a |
| Descriptive data | 14* | (a) Give characteristics of study participants (eg demographic, clinical, social) and information on exposures and potential confounders | 12 |
|  |  | (b) Indicate number of participants with missing data for each variable of interest | n/a |
| Outcome data | 15* | Report numbers of outcome events or summary measures | 13-14 |
| Main results | 16 | (*a*) Give unadjusted estimates and, if applicable, confounder-adjusted estimates and their precision (eg, 95% confidence interval). Make clear which confounders were adjusted for and why they were included | 14-17 |
|  |  | (*b*) Report category boundaries when continuous variables were categorized | 14-17 |
|  |  | (*c*) If relevant, consider translating estimates of relative risk into absolute risk for a meaningful time period | n/a |
| Other analyses | 17 | Report other analyses done—eg analyses of subgroups and interactions, and sensitivity analyses | 14-17 |
| Discussion | | |  |
| Key results | 18 | Summarise key results with reference to study objectives | 18 |
| Limitations | 19 | Discuss limitations of the study, taking into account sources of potential bias or imprecision. Discuss both direction and magnitude of any potential bias | 22 |
| Interpretation | 20 | Give a cautious overall interpretation of results considering objectives, limitations, multiplicity of analyses, results from similar studies, and other relevant evidence | 19-22 |
| Generalisability | 21 | Discuss the generalisability (external validity) of the study results | 22 |
| Other information | | |  |
| Funding | 22 | Give the source of funding and the role of the funders for the present study and, if applicable, for the original study on which the present article is based | 1 |

**Supplementary Table 8.** STROBE Statement—Checklist of items that should be included in reports of *cross-sectional studies.*
